# Comparative effectiveness of two- and three-dose schedules involving AZD1222 and BNT162b2 in people with kidney disease: a linked OpenSAFELY and UK Renal Registry cohort study

**DOI:** 10.1101/2022.11.16.22282396

**Authors:** The OpenSAFELY Collaborative, Edward PK Parker, Elsie MF Horne, William J Hulme, John Tazare, Bang Zheng, Edward J Carr, Fiona Loud, Susan Lyon, Viyaasan Mahalingasivam, Brian MacKenna, Amir Mehrkar, Miranda Scanlon, Shalini Santhakumaran, Retha Steenkamp, Ben Goldacre, Jonathan AC Sterne, Dorothea Nitsch, Laurie A Tomlinson, The LH&W NCS (or CONVALESCENCE) Collaborative

**Affiliations:** London School of Hygiene and Tropical Medicine, Keppel Street, London, WC1E 7HT, UK; Population Health Sciences, University of Bristol, Oakfield House, Oakfield Grove, Bristol, BS8 2BN, UK; NIHR Bristol Biomedical Research Centre, Bristol, UK; Bennett Institute for Applied Data Science, Nuffield Department of Primary Care Health Sciences, University of Oxford, OX2 6GG, UK; The Francis Crick Institute, London, NW1 1AT, UK; Kidney Care UK, Alton, UK; Patient Council, UK Kidney Association, Bristol, UK; Kidney Research UK, Peterborough, UK; UK Renal Registry, Bristol, UK; Health Data Research UK South-West, Bristol, UK

**Keywords:** COVID-19, chronic kidney disease, NHS England, SARS-CoV-2, vaccination, effectiveness

## Abstract

**Background:** Kidney disease is a key risk factor for COVID-19-related mortality and suboptimal vaccine response. Optimising vaccination strategies is essential to reduce the disease burden in this vulnerable population.

**Methods:** With the approval of NHS England, we performed a retrospective cohort study to estimate the comparative effectiveness of schedules involving AZD1222 (AZ; ChAdOx1-S) and BNT162b2 (BNT) among people with kidney disease. Using linked primary care and UK Renal Registry records in the OpenSAFELY-TPP platform, we identified adults with stage 3– 5 chronic kidney disease, dialysis recipients, and kidney transplant recipients. We used Cox proportional hazards models to compare COVID-19-related outcomes and non-COVID-19 death after two-dose (AZ–AZ vs BNT–BNT) and three-dose (AZ–AZ–BNT vs BNT–BNT– BNT) schedules.

**Findings:** After two doses, incidence during the Delta wave was higher in AZ–AZ (n=257,580) than BNT–BNT recipients (n=169,205; adjusted hazard ratios [95% CIs] 1·43 [1·37–1·50], 1·59 [1·43–1·77], 1·44 [1·12–1·85], and 1·09 [1·02–1·17] for SARS-CoV-2 infection, COVID-19-related hospitalisation, COVID-19-related death, and non-COVID-19 death, respectively). Findings were consistent across disease subgroups, including dialysis and transplant recipients. After three doses, there was little evidence of differences between AZ– AZ–BNT (n=220,330) and BNT–BNT–BNT recipients (n=157,065) for any outcome during a period of Omicron dominance.

**Interpretation:** Among individuals with moderate-to-severe kidney disease, two doses of BNT conferred stronger protection than AZ against SARS-CoV-2 infection and severe disease. A subsequent BNT dose levelled the playing field, emphasising the value of heterologous RNA doses in vulnerable populations.

**Funding:** National Core Studies, Wellcome Trust, MRC, and Health Data Research UK.

**Research in context:** *Evidence before this study:* We searched Medline for studies published between 1^st^ December 2020 and 7^th^ September 2022 using the following term: *“(coronavir* or covid* or sars*) and (vaccin* or immunis* or immuniz*) and (kidney or dialysis or h?emodialysis or transplant or renal) and (efficacy or effectiveness)”*. We identified studies reporting on the effectiveness of various COVID-19 vaccines in individuals with chronic kidney disease (CKD) or end-stage renal disease. Several studies have reported no clear differences in effectiveness against outcomes of varying severity after two doses of BNT162b2 or AZD1222 compared to unvaccinated controls, which is contrary to the significantly higher antibody levels observed after BNT162b2 in immunogenicity studies. One study also showed that a third dose of RNA vaccine restored some protection against the Omicron variant among BNT162b2- and AZD1222-primed individuals, with no clear differences between these groups. This finding is consistent with immunogenicity data suggesting that a third dose of BNT162b2 may reduce the gap in antibody levels observed after two of AZD1222 versus BNT162b2. Notably, we found few studies directly comparing effectiveness in BNT162b2 versus AZD1222 recipients, which reduces biases associated with comparison to a small and potentially unrepresentative group of unvaccinated controls. We also found no studies exploring COVID-19 vaccine effectiveness in kidney disease groups of varying severity (CKD, dialysis, and kidney transplant).

*Added value of this study:* This is the largest study to compare the effectiveness of two- and three-dose regimens involving AZD1222 and BNT162b2 among people with moderate-to-severe kidney disease. We compared effectiveness after two and three doses in 426,780 and 377,395 individuals, respectively, and harnessed unique data linkages between primary care records and UK Renal Registry data to identify people with CKD and end-stage renal disease (including dialysis and kidney transplant recipients) with high accuracy. During the Delta wave of infection, we observed a higher risk of COVID-19-related outcomes of varying severity after two doses of AZD1222 versus BNT162b2, with consistent findings in CKD, dialysis, and transplant subgroups. After a third dose of BNT162b2, AZD1222- and BNT162b2-primed individuals had similar rates of COVID-19-related outcomes during a period of Omicron dominance. Implications of all the available evidence A growing body of immunogenicity and effectiveness data – including the present study – suggest that two doses of BNT162b2 confers stronger protection than AZD1222 among people with moderate-to-severe kidney disease. However, a third dose of BNT162b2 appears to compensate for this immunity deficit, providing equivalent protection in BNT162b2- and AZD1222-primed individuals. Achieving high coverage with additional RNA vaccine doses (whether homologous or heterologous) has the capacity to reduce the burden of disease in this vulnerable population.

## Background

During the SARS-CoV-2 pandemic, the burden of morbidity and mortality has not been shared equally across society. Social and demographic factors have underpinned stark inequities in SARS-CoV-2 exposure, while clinical factors have shaped the ensuing risk of harm among those infected. Although COVID-19 vaccines have the potential to mitigate these inequities, people with compromised immune systems may fail to mount a protective response to primary or booster doses^1,2^, leaving them at increased risk of subsequent infection and disease compared with healthy adults^3,4^.

Kidney disease is a key risk factor for both COVID-19-related mortality and suboptimal COVID-19 vaccine response^4,5^. Observational studies of vaccine immunogenicity have highlighted possible ways to optimise immunisation strategies in this population. In people receiving haemodialysis, antibody levels are significantly lower following two doses of the vectored vaccine AZD1222 (AZ; ChAdOx1-S) compared with the RNA vaccine BNT162b2 (BNT)^6,7^. However, a heterologous BNT third dose among AZ-primed individuals (AZ–AZ– BNT) appears to reduce the immunogenicity gap, inducing antibody levels equivalent^7^ or closer^8,9^ to those observed after a homologous three-dose series (BNT–BNT–BNT).

The extent to which these immunogenicity data translate to protection against infection and severe COVID-19 remains uncertain. To address this, we harnessed unique data linkages within the OpenSAFELY-TPP database to estimate the comparative effectiveness of two-dose (AZ–AZ vs BNT–BNT) and three-dose (AZ–AZ–BNT vs BNT–BNT–BNT) schedules among people with kidney disease in England.

## Methods

### Data sources

All data were linked, stored, and analysed securely within the OpenSAFELY platform https://opensafely.org/. OpenSAFELY is a data analytics platform created by our team on behalf of NHS England to address urgent COVID-19 research questions. The dataset analysed within OpenSAFELY-TPP is based on 24 million people currently registered with GP surgeries using TPP SystmOne software. Data include pseudonymised data such as coded diagnoses, medications, and physiological parameters. No free text data are included. All code is shared openly for review and re-use under MIT open license (https://github.com/opensafely/ckd-coverage-ve). Detailed pseudonymised patient data is potentially re-identifiable and therefore not shared. Primary care data are linked through OpenSAFELY with other pseudonymised datasets, including COVID-19 testing records via the Second Generation Surveillance System (SGSS), A&E attendance and hospital records via NHS Digital’s Hospital Episode Statistics (HES), national death registry records from the Office for National Statistics (ONS), and renal replacement therapy (RRT) status via the UK Renal Registry (UKRR). Vaccination status is directly available within GP records via the National Immunisation Management System.

### Study population

We defined separate cohorts for the two- and three-dose analyses. Baseline characteristics were defined as of dose 1 (two-dose cohort) or dose 3 (three-dose cohort), except for age, which was calculated as of 31^st^ March 2021 as per UK Health Security Agency recommendations^10^. We assessed potential eligibility among individuals who were at least 16 years of age and had been registered in OpenSAFELY-TPP for at least 3 months before their first COVID-19 vaccine dose.

Each cohort comprised: (i) individuals receiving RRT (dialysis or transplant), as listed within the UKRR as of 31^st^ December 2020; and (ii) individuals with evidence of stage 3–5 chronic kidney disease (CKD) in the absence of RRT. Individuals with CKD were identified based on their most recent serum creatinine measurement in the 2 years preceding baseline. Creatinine levels were converted into estimated glomerular filtration rate (eGFR) using the CKD epidemiology collaboration equation without specification of ethnicity^11^, and an eGFR of <60 ml/min/1·73 m^2^ used as a threshold for inclusion. To assess CKD severity, we distinguished between stage 3a (eGFR of 45–59 ml/min/1·73 m^2^), stage 3b (eGFR of 30–44 ml/min/1·73 m^2^), and stage 4–5 CKD (eGFR <30 ml/min/1·73 m^2^). Individuals with primary care codes suggesting prior dialysis or kidney transplant but absent from the UKRR were excluded given their ambiguous kidney disease status at the point of recruitment.

Individuals were included in the two-dose cohort if they: (i) had complete data on sex, ethnicity, NHS region, and index of multiple deprivation (IMD); (ii) received AZ–AZ or BNT–BNT; (iii) received their first vaccine dose on or after 4^th^ January 2021 (when both AZ and BNT were in concurrent use); (iv) had a dose 1–2 interval of 8–14 weeks; (v) were not health or social care workers, residents in care or nursing homes, medically housebound, or receiving end-of-life care (given atypical testing and exposure patterns in these groups); (vi) were not aged ≥80 years (given significant vaccination in this age group before AZ was available); (vii) received their second dose on or before 17^th^ October 2021 (ensuring 30 days of potential follow-up); and (viii) had no documented SARS-CoV-2 infection in the 90 days preceding dose 1 or between doses 1 and 2.

Individuals were included in the three-dose cohort if they fulfilled criteria (i) to (vi) above and received BNT as a third dose between 1^st^ September 2021 (the date on which third primary doses were recommended for immunosuppressed individuals^12^) and 1^st^ March 2022. Additionally, individuals were excluded if they had a dose 2–3 interval of <12 weeks or were infected with SARS-CoV-2 between doses 1 and 3 (to mitigate the influence of hybrid immunity). We did not distinguish between third primary and booster doses.

### Outcomes

We assessed the following post-vaccination outcomes: SARS-CoV-2 infection; COVID-19-related hospitalisation; COVID-19-related death; and non-COVID-19 death (see **Supplementary Table S1** for coding details). For the two-dose cohort, follow-up started on the date of dose 2 and extended until 16^th^ November 2021 (2 months after all individuals became eligible for a third dose^13^). For the three-dose cohort, follow-up started on the date of dose 3 and extended until 31^st^ March 2022 for SARS-CoV-2 infection (the date when free testing for the public came to an end) or 21^st^ May 2022 for other outcomes (2 months after the launch of the spring booster campaign^14,15^).

### Potential confounding variables

We defined the following potential confounders in each cohort: age; sex; ethnicity; social deprivation based on IMD quintile; setting (urban, urban conurbation, or rural,); kidney disease subgroup (CKD3a, CKD3b, CKD4–5, dialysis, or transplant); clinical comorbidities that influenced vaccine prioritisation (**Supplementary Table S1**); prior SARS-CoV-2 infection; and number of SARS-CoV-2 (0, 1, 2, or 3+) in the 90 days preceding 4^th^ January 2021 (two-dose cohort) or 1^st^ September 2021 (three-dose cohort) as an indicator of testing behaviour. After accounting for exclusions relating to incomplete demographic data (see above), there were no missing values as remaining variables were defined by the presence or absence of codes or events.

### Statistical analysis

Follow-up started on the day of vaccination (dose 2 or 3) and was censored at the earliest of death, deregistration, the administration of a subsequent COVID-19 vaccine dose, 182 days, the end of the study period (as defined above), or the outcome of interest. We used the Kaplan-Meier (KM) method to estimate cumulative incidence and associated 95% confidence intervals (CIs). Risk ratios (RRs) were estimated based on KM estimates, alongside 95% CIs derived from the sum of squares of the log-KM standard errors. We used Cox proportional hazards models to compare effectiveness of AZ–AZ versus BNT–BNT (two-dose cohort) and AZ–AZ–BNT versus BNT–BNT–BNT (three-dose cohort) for each outcome. Hazard ratios (HRs) and 95% CIs were estimated for the whole study period and during the following time periods: days 1–14, 14–70, 71–126, and 127–182.

For each outcome, we fitted: (i) unadjusted models; (ii) models stratified by NHS region and adjusting for calendar-time effects by including a natural cubic spline for the date of vaccination with two knots at the first and second tertiles; and (iii) fully adjusted models additionally adjusting for the demographic and clinical confounders described above (with age as a quadratic polynomial). Fully adjusted models for the overall study period were explored in the following subgroups: CKD3, CKD4–5, kidney transplant, dialysis, and any RRT (combining transplant and dialysis). To avoid issues with model convergence, we excluded binary covariates if cross-tabulating the variable with vaccine group yielded any cell with fewer than three outcome events. For categorical variables with more than two levels, we merged categories until all levels fulfilled these cross-tabulation criteria or one level remained, in which case the variable was excluded. Final model structures are summarised in **Supplementary Tables S2** and **S3**.

As an alternative to confounder adjustment via regression, we conducted a sensitivity analysis in which vaccination groups were matched 1:1 based on: age (within 3 years); date of vaccination (within 3 days); date of preceding dose (within 7 and 14 days for the two- and three-dose cohorts, respectively); sex; IMD quintile; NHS region; kidney disease subgroup; classification as clinically extremely vulnerable; prior SARS-CoV-2 infection; and the presence of any indicator of immunosuppression (recent immunosuppressive therapy, permanent immunosuppression, asplenia, haematologic malignancy, or transplant).

In compliance with re-identification minimisation requirements for OpenSAFELY, we rounded any reported counts to the nearest 5, redacted non-zero counts of ≤10, and delayed KM steps until ≥5 events had occurred.

### Patient and public involvement

People living with kidney disease were among the team that planned this work and reviewed the manuscript. The work will be shared through involvement of the team with kidney charities and other organisations.

## Results

### Study population

426,785 individuals with kidney disease were eligible for the two-dose cohort (**Supplementary Table S4**), of whom 257,580 (60%) received AZ–AZ and 169,205 (40%) received BNT–BNT. Baseline clinical and demographic characteristics were similar across the vaccine groups (**Table 1**). The most common kidney disease subgroup was CKD3a (76% vs 75% among AZ–AZ vs BNT–BNT recipients, respectively), followed by CKD3b (17% vs 18%), CKD4–5 (4% vs 4%), transplant (2% vs 2%), and dialysis (1% vs 1%).

**Table 1.** Baseline characteristics of two-dose vaccination cohort.

| Characteristic | Unmatched |  | Matched |  |
| --- | --- | --- | --- | --- |
|  | AZ-AZ<br>N = 257,580 | BNT-BNT<br>N = 169,205 | AZ-AZ<br>N = 130,765 | BNT-BNT<br>N = 130,65 |
| **Age** |  |  |  |  |
| 16–64 | 54,330 (21.1%) | 29,485 (17.4%) | 15,040 (11.5%) | 18,015 (13.8%) |
| 65–69 | 41,890 (16.3%) | 21,740 (12.8%) | 19,120 (14.6%) | 19,030 (14.6%) |
| 70–74 | 80,215 (31.1%) | 45,125 (26.7%) | 43,710 (33.4%) | 39,420 (30.1%) |
| 75–79 | 81,150 (31.5%) | 72,855 (43.1%) | 52,895 (40.5%) | 54,300 (41.5%) |
| **Sex** |  |  |  |  |
| Female | 136,300 (52.9%) | 89,095 (52.7%) | 69,475 (53.1%) | 69,475 (53.1%) |
| Male | 121,280 (47.1%) | 80,110 (47.3%) | 61,295 (46.9%) | 61,295 (46.9%) |
| **Ethnicity** |  |  |  |  |
| White | 240,905 (93.5%) | 159,055 (94.0%) | 124,655 (95.3%) | 124,810 (95.4%) |
| Black | 4,675 (1.8%) | 2,535 (1.5%) | 1,330 (1.0%) | 1,370 (1.0%) |
| South Asian | 8,825 (3.4%) | 5,685 (3.4%) | 3,555 (2.7%) | 3,360 (2.6%) |
| Mixed | 1,370 (0.5%) | 835 (0.5%) | 475 (0.4%) | 530 (0.4%) |
| Other | 1,805 (0.7%) | 1,095 (0.6%) | 750 (0.6%) | 695 (0.5%) |
| **Index of multiple deprivation quintile** |  |  |  |  |
| 1 most deprived | 43,860 (17.0%) | 27,650 (16.3%) | 20,270 (15.5%) | 20,270 (15.5%) |
| 2 | 49,590 (19.3%) | 32,590 (19.3%) | 24,695 (18.9%) | 24,695 (18.9%) |
| 3 | 58,295 (22.6%) | 38,860 (23.0%) | 30,420 (23.3%) | 30,420 (23.3%) |
| 4 | 55,655 (21.6%) | 36,980 (21.9%) | 29,130 (22.3%) | 29,130 (22.3%) |
| 5 least deprived | 50,175 (19.5%) | 33,120 (19.6%) | 26,250 (20.1%) | 26,250 (20.1%) |
| **Setting** |  |  |  |  |
| Urban city or town | 138,500 (53.8%) | 90,055 (53.2%) | 71,810 (54.9%) | 70,565 (54.0%) |
| Urban conurbation | 50,345 (19.5%) | 33,035 (19.5%) | 23,815 (18.2%) | 23,345 (17.9%) |
| Rural | 68,735 (26.7%) | 46,115 (27.3%) | 35,145 (26.9%) | 36,855 (28.2%) |
| **Kidney disease** |  |  |  |  |
| CKD3a | 195,125 (75.8%) | 126,335 (74.7%) | 102,350 (78.3%) | 102,350 (78.3%) |
| CKD3b | 44,255 (17.2%) | 30,630 (18.1%) | 22,365 (17.1%) | 22,365 (17.1%) |
| CKD4–5 | 10,325 (4.0%) | 6,985 (4.1%) | 3,825 (2.9%) | 3,825 (2.9%) |
| RRT (dialysis) | 2,335 (0.9%) | 1,870 (1.1%) | 505 (0.4%) | 505 (0.4%) |
| RRT (transplant) | 5,540 (2.2%) | 3,385 (2.0%) | 1,720 (1.3%) | 1,720 (1.3%) |
| **Primary care coding of kidney disease** |  |  |  |  |
| CKD3–5 | 137,395 (53.3%) | 96,025 (56.8%) | 72,400 (55.4%) | 73,350 (56.1%) |
| Dialysis code | 5,470 (2.1%) | 3,655 (2.2%) | 1,520 (1.2%) | 1,525 (1.2%) |
| Kidney transplant code | 5,800 (2.3%) | 3,585 (2.1%) | 1,740 (1.3%) | 1,725 (1.3%) |
| **Morbidities** |  |  |  |  |
| Immunosuppression | 16,155 (6.3%) | 10,885 (6.4%) | 6,345 (4.9%) | 6,360 (4.9%) |
| Severe obesity | 15,815 (6.1%) | 10,165 (6.0%) | 7,665 (5.9%) | 7,760 (5.9%) |
| Diabetes | 70,690 (27.4%) | 47,975 (28.4%) | 37,580 (28.7%) | 36,390 (27.8%) |
| Chronic respiratory disease (inc. asthma) | 27,565 (10.7%) | 18,585 (11.0%) | 14,485 (11.1%) | 14,010 (10.7%) |
| Chronic heart disease | 88,375 (34.3%) | 61,725 (36.5%) | 48,160 (36.8%) | 47,685 (36.5%) |
| Chronic liver disease | 11,165 (4.3%) | 7,165 (4.2%) | 5,595 (4.3%) | 5,350 (4.1%) |
| Asplenia | 2,490 (1.0%) | 1,580 (0.9%) | 985 (0.8%) | 930 (0.7%) |
| Haematologic cancer | 4,675 (1.8%) | 3,355 (2.0%) | 1,985 (1.5%) | 1,970 (1.5%) |
| Organ transplant (non-kidney) | 960 (0.4%) | 525 (0.3%) | 290 (0.2%) | 260 (0.2%) |
| Chronic neurological disease | 26,335 (10.2%) | 17,920 (10.6%) | 14,390 (11.0%) | 13,775 (10.5%) |
| Learning disability | 1,140 (0.4%) | 605 (0.4%) | 375 (0.3%) | 400 (0.3%) |
| Severe mental illness | 3,785 (1.5%) | 2,135 (1.3%) | 1,650 (1.3%) | 1,605 (1.2%) |
| Clinically extremely vulnerable | 48,735 (18.9%) | 32,475 (19.2%) | 20,485 (15.7%) | 20,485 (15.7%) |
| **Prior SARS-CoV-2 infection** | 3,880 (1.5%) | 2,235 (1.3%) | 565 (0.4%) | 565 (0.4%) |
| **No. of SARS-CoV-2 tests in 90-day pre-vaccination window** |  |  |  |  |
| 0 | 221,940 (86.2%) | 145,815 (86.2%) | 114,135 (87.3%) | 114,665 (87.7%) |
| 1 | 22,910 (8.9%) | 15,020 (8.9%) | 11,010 (8.4%) | 10,985 (8.4%) |
| 2 | 6,095 (2.4%) | 4,005 (2.4%) | 2,845 (2.2%) | 2,735 (2.1%) |
| 3+ | 6,630 (2.6%) | 4,360 (2.6%) | 2,780 (2.1%) | 2,380 (1.8%) |
| **Region** |  |  |  |  |
| East of England | 55,955 (21.7%) | 38,525 (22.8%) | 28,340 (21.7%) | 28,340 (21.7%) |
| Midlands | 62,190 (24.1%) | 40,340 (23.8%) | 33,535 (25.6%) | 33,535 (25.6%) |
| London | 5,590 (2.2%) | 5,715 (3.4%) | 2,650 (2.0%) | 2,650 (2.0%) |
| North East and Yorkshire | 48,920 (19.0%) | 30,425 (18.0%) | 23,490 (18.0%) | 23,490 (18.0%) |
| North West | 24,590 (9.5%) | 14,535 (8.6%) | 11,005 (8.4%) | 11,005 (8.4%) |
| South East | 13,740 (5.3%) | 9,020 (5.3%) | 6,540 (5.0%) | 6,540 (5.0%) |
| South West | 46,600 (18.1%) | 30,640 (18.1%) | 25,205 (19.3%) | 25,205 (19.3%) |
| **JCVI priority group** |  |  |  |  |
| 3 (75+) | 81,150 (31.5%) | 72,855 (43.1%) | 52,895 (40.5%) | 54,300 (41.5%) |
| 4 (70+ or clinically extremely vulnerable) | 103,575 (40.2%) | 58,180 (34.4%) | 50,920 (38.9%) | 46,700 (35.7%) |
| 5 (65+) | 34,040 (13.2%) | 17,635 (10.4%) | 16,195 (12.4%) | 16,095 (12.3%) |
| 6 (16–65 and clinically vulnerable) | 38,820 (15.1%) | 20,535 (12.1%) | 10,755 (8.2%) | 13,665 (10.5%) |

377,395 individuals were eligible for the three-dose cohort, of whom 222,330 (58%) received AZ–AZ–BNT and 157,065 (42%) received BNT–BNT–BNT. Again, baseline clinical and demographic characteristics were similar across vaccine groups (**Supplementary Table S5**). When stratified by kidney disease subgroup, baseline characteristics remained similar across vaccine groups (**Supplementary Tables S6** and **S7**). RRT recipients were notably younger than people with CKD and had a higher prevalence of immunosuppression, non-White ethnicity, and prior SARS-CoV-2 infection.

### Comparative effectiveness after two doses

Eligible individuals began to receive their second doses in March 2021, with follow-up spanning the Delta wave of SARS-CoV-2 up to the analysis cut-off of 16^th^ November 2021 (**Supplementary Figure S1A**). There were a total of 10,405 SARS-CoV-2 infections, 1,660 COVID-19-related hospitalisations, 305 COVID-19-related deaths, and 4,045 non-COVID-19 deaths across a median follow-up time of 182 (interquartile range [IQR] 182–182) days for each outcome (**Figure 1**).

**Figure 1.**
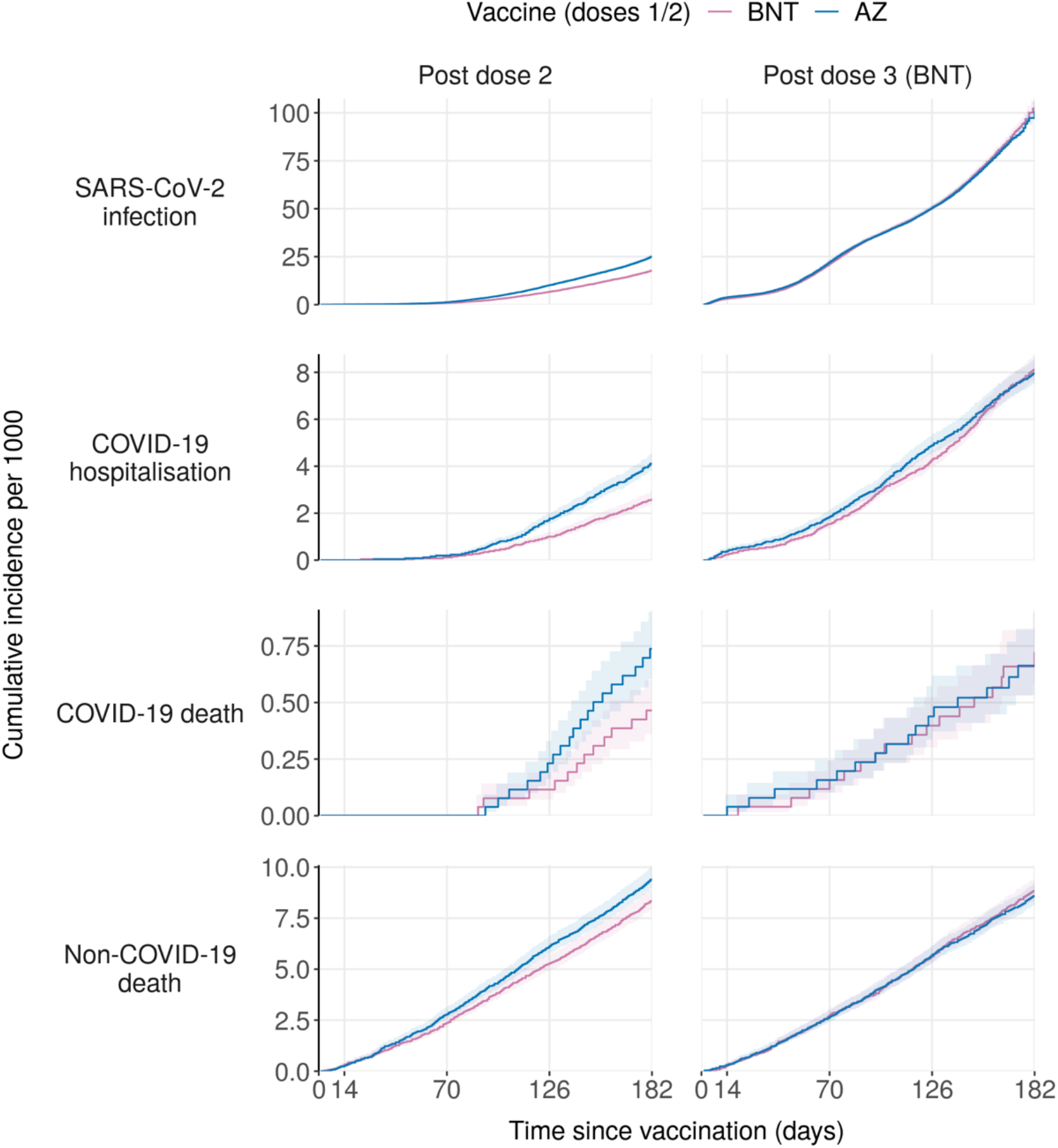
Cumulative incidence rates by vaccine group. Kaplan-Meier estimates of cumulative incidence in matched two-dose and three-dose sub-cohorts. Kaplan-Meier steps are delayed until ≥5 events occur in compliance with re-identification minimisation requirements in OpenSAFELY. Data for the matched sub-cohort are shown here to minimise confounding associated with key matching variables; see **Supplementary Figure S1** for equivalent plots relating to the unmatched cohorts. Numbers at risk at days 0, 14, 70, and 126 are provided in **Supplementary Tables S8** (two-dose cohort) and **S10** (three-dose cohort). AZ, AZD1222 (AstraZeneca); BNT, BNT162b2 (Pfizer/BioNTech).

Incidence rates and HRs are summarised in **Figures 1**–**3** and **Supplementary Table S8**. Across the overall follow-up period, SARS-CoV-2 infections occurred at a rate of 58·7 per 1000 person-years in AZ–AZ recipients and 37·1 per 1000 person-years in BNT–BNT recipients (fully adjusted HR [95% CI] of 1·43 [1·37–1·50]). HRs were similar across modelling strategies, including in the matched sub-cohort (**Supplementary Table S8)**. When stratified by time period, discrepancies between AZ–AZ and BNT–BNT recipients were absent from days 1–14 (when few cases occurred), but consistent across all subsequent time periods (**Figure 3**).

**Figure 3.**
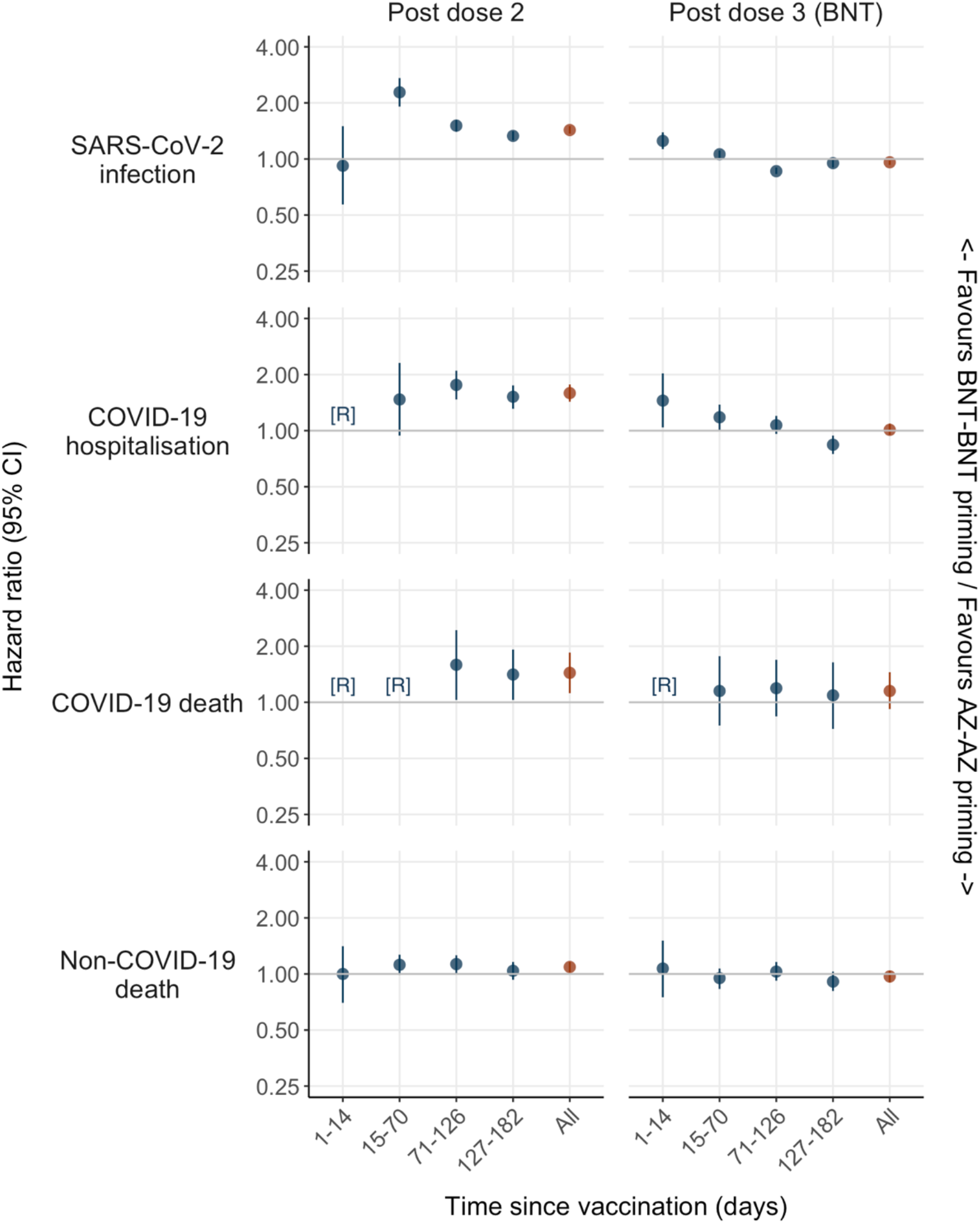
Hazard ratios for the comparative effectiveness of two- and three-dose schedules. See **Supplementary Tables S8** (two-dose cohort) and **S8** (three-dose cohort) for associated population sizes and incidence rates. AZ, AZD1222 (AstraZeneca); BNT, BNT162b2 (Pfizer/BioNTech); CI, confidence interval; [R], redacted due to low event counts in one or more groups.

Similar discrepancies between AZ–AZ and BNT–BNT recipients were observed for more severe outcomes. Overall incidence rates for COVID-19-related hospitalisation were 9·4 per 1000 person-years in AZ–AZ recipients and 5·7 per 1000 person-years in BNT–BNT recipients (fully adjusted HR 1·59 [1·43–1·77]). COVID-19-related deaths occurred at a rate of 1·7 per 1000 person-years in AZ–AZ recipients and 1·1 per 1000 person-years in BNT–BNT recipients (fully adjusted HR 1·44 [1·12–1·85]). Where event counts were sufficient to enable estimation, discrepancies for both outcomes were consistent across time periods and modelling strategies, including in the matched sub-cohort (**Supplementary Table S8**). Non-COVID-19 deaths occurred at a marginally higher rate in AZ–AZ than BNT–BNT recipients (fully adjusted HR 1·09 [1·02–1·16]).

COVID-19-related outcomes were consistently more common in RRT recipients than people with CKD3 (**Supplementary Table S9**), with intermediate incidence among people with CKD4–5. For SARS-CoV-2 infection and COVID-19-related hospitalisation, comparative effectiveness estimates were highly consistent across kidney disease subgroups, including transplant and dialysis recipients (HRs of 1·2–1·7 in favour of BNT–BNT; **Figure 2**). Subgroup-specific estimates for COVID-19-related deaths were generally non-significant (i.e., CIs crossed 1), albeit constrained by low event counts in individual subgroups. The discrepancy in non-COVID-19 deaths observed for the overall cohort appears to be driven by individuals with CKD3 (fully adjusted HR 1·11 [1·03–1·19]); no significant difference was observed for people with CKD4–5 or in RRT recipients (in contrast to the significant discrepancies in COVID-19-related outcomes in these subgroups).

**Figure 2.**
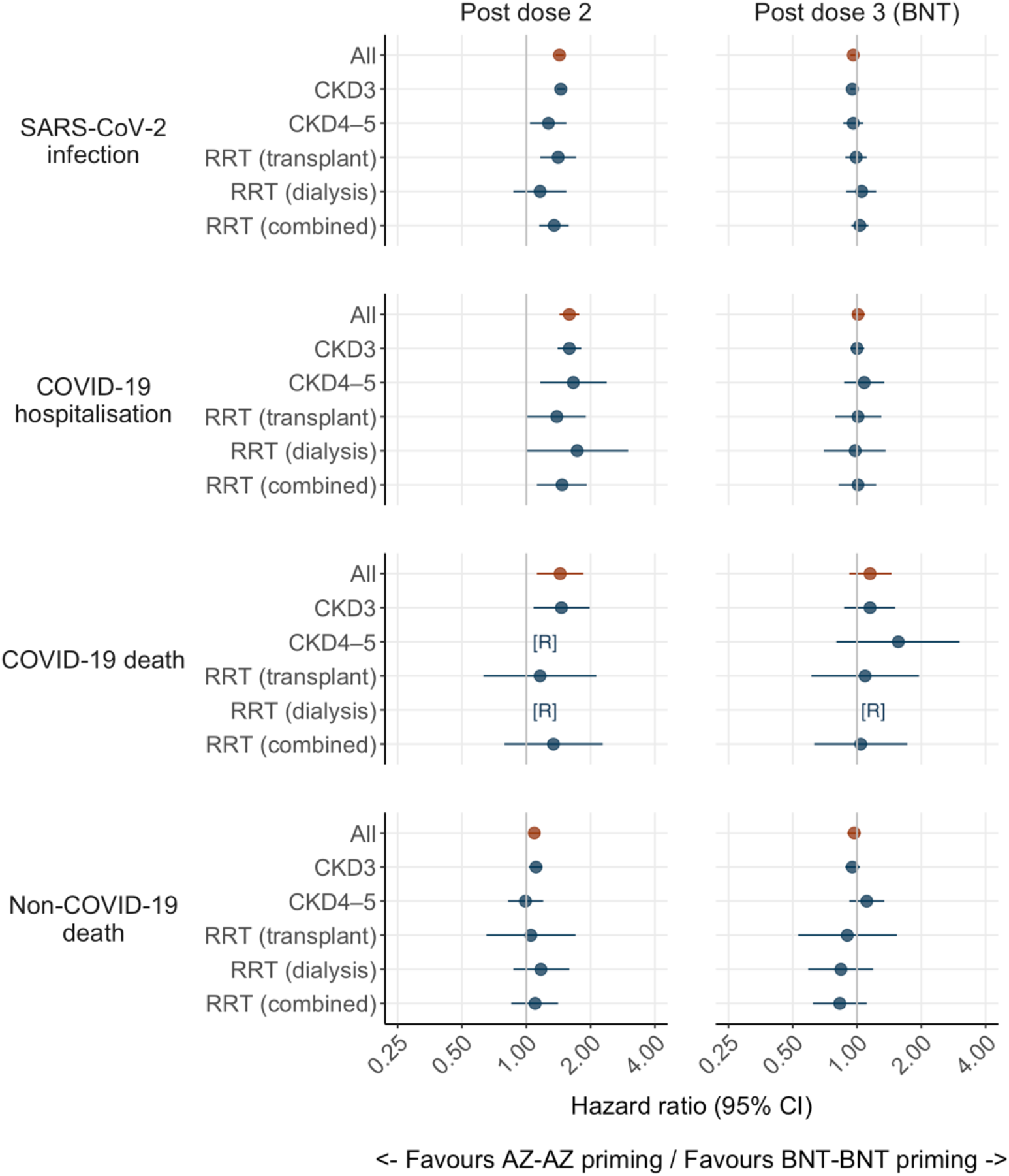
Hazard ratios for the comparative effectiveness of two- and three-dose schedules in kidney disease subgroups. See **Supplementary Tables S7** (two-dose cohort) and **S9** (three-dose cohort) for associated population sizes and incidence rates. AZ, AZD1222 (AstraZeneca); BNT, BNT162b2 (Pfizer/BioNTech); CI, confidence interval; CKD, chronic kidney disease; [R], redacted due to low event counts in one or both vaccine groups; RRT, renal replacement therapy.

### Comparative effectiveness after BNT162b2 third dose

Follow-up for the three-dose cohort spanned periods of Delta and Omicron dominance between September 2021 and May 2022 (**Supplementary Figure S1B**). There were 27,320 SARS-CoV-2 infections, 3,360 COVID-19-related hospitalisations, 325 COVID-19-related deaths, and 3,420 non-COVID-19 deaths across a median follow-up time of 146 [IQR 127– 160], 182 [170–182], 182 [170–182], and 182 [170–182] days, respectively (**Figure 1**). A total of 25,105 (92%) SARS-CoV-2 infections occurred on or after 15^th^ December 2021, when Omicron became the dominant variant in England. The proportion of SARS-CoV-2 infections occurring during the Omicron era rose across time periods from 330/1,535 (21%) in days 1– 14 to 6,940/7,915 (88%) in days 15–70 and >99% thereafter.

Full model outputs for the three-dose cohort are provided in **Supplementary Table S10**. Across the overall follow-up period, SARS-CoV-2 infections occurred at a similar rate in AZ–AZ–BNT versus BNT–BNT–BNT recipients (156·2 vs 166·4 per 1000 person-years, respectively; fully adjusted HR 0·96 [0·93–0·98]; **Figures 1** and **2**). When broken down by time period, infections initially occurred at a higher rate in AZ–AZ–BNT recipients (fully adjusted HRs of 1·25 [1·13–1·39] and 1·06 [1·01–1·11] for days 1–14 and 15–70, respectively), while the inverse was true from day 71 onwards, resulting in approximate parity across the overall study period. Findings were similar across modelling strategies, supporting equivalent protection for AZ–AZ–BNT versus BNT–BNT–BNT across the study period (e.g., HRs of 0·98 [0·96–1·02] for the matched sub-cohort; **Supplementary Table S10**).

COVID-19-related hospitalisations occurred at a rate of 19·1 per 1000 person-years in AZ– AZ–BNT recipients and 19·0 per 1000 person-years in BNT–BNT–BNT recipients (fully adjusted HR 1.01 [0·95–1·09]). As observed for SARS-CoV-2 infection, hospitalisations were initially more common among AZ–AZ–BNT recipients, but reached parity across the overall study period (**Figures 1** and **3**). COVID-19-related deaths occurred at a rate of 1·9 per 1000 person-years in AZ–AZ–BNT recipients and 1·8 per 1000 person-years in BNT–BNT–BNT recipients (fully adjusted HR 1·15 [0·92–1·45]). Likewise, non-COVID-19 deaths occurred at a similar rate across the two vaccine groups (fully adjusted HR 0·97 [0·90–1·04]). When analyses were stratified by kidney disease subgroup, we observed no clinically important differences in event rates between vaccine groups for any outcome in any subgroup (**Figure 2** and **Supplementary Table S11**), including transplant and dialysis recipients.

## Discussion

Optimising the use of available COVID-19 vaccines among people with kidney disease is crucial to reduce the burden of morbidity and mortality in this vulnerable population. The present study suggests that a two-dose series of BNT conferred stronger protection than AZ against SARS-CoV-2 infection, COVID-19-related hospitalisation, and COVID-19-related death among people with kidney disease in England during the Delta wave of the pandemic. These findings were consistent across CKD3, CKD4–5, transplant and dialysis subgroups. By contrast, after administration of BNT as a third dose, AZ- and BNT-primed individuals exhibited similar risks of COVID-19 during a period dominated by the Omicron variant.

Our findings are consistent with previous immunology studies that reported higher antibody levels following BNT–BNT versus AZ–AZ^16-18^, including studies in dialysis and kidney transplant recipients^6,19^. Higher effectiveness for BNT–BNT versus AZ–AZ has also been reported during the Delta wave in study populations not restricted to individuals with kidney disease^20,21^. One study of solid organ transplant recipients in England showed no reduction in SARS-CoV-2 infection rates relative to unvaccinated individuals following AZ– AZ or BNT–BNT, and a reduction in post-infection mortality for AZ–AZ but not BNT– BNT^22^. In addition, previous cohort studies focusing on people receiving haemodialysis have documented no clear differences in effectiveness or post-infection progression between BNT–BNT and AZ–AZ^8,23^. By contrast, we found that people receiving RRT (dialysis or kidney transplant) were less likely to experience SARS-CoV-2 infection and COVID-19-related hospitalisation following BNT–BNT versus AZ–AZ. In contrast to previous studies, we limited enrolment to periods of concomitant AZ and BNT usage. We also harnessed primary care data linkages to identify detailed comorbidity data and reduce confounding of comparative effectiveness estimates.

The administration of BNT as a third dose appeared to compensate for the discrepancies in protection between AZ- and BNT-primed individuals. This finding is consistent with observational and clinical studies of third-dose immunogenicity in healthy adults^16,24^ and individuals with kidney disease^7,9,25^, as well as vaccine effectiveness data from the general population^26^. One previous study explored short-term vaccine effectiveness of BNT as a third dose following AZ or BNT priming among haemodialysis recipients, reporting increased protection against Omicron infection for boosted compared with unboosted individuals and no significant differences according to primary vaccine group^27^. We show the equivalent effectiveness of AZ–AZ–BNT versus BNT–BNT–BNT to persist for up to 182 days against infection, COVID-19-related hospitalisation, and COVID-19-related death. Interestingly, the higher rates of SARS-CoV-2 infection and COVID-19-related hospitalisation observed after AZ–AZ versus BNT–BNT persisted for days 1–14 after the third dose of BNT, consistent with a lag of 1–2 weeks while the additional dose takes effect.

Our study is strengthened by the scale and representativeness of the OpenSAFELY-TPP database^28^, offering the statistical power to explore endpoints such as COVID-19-related hospitalisation and death. The unique use of a gold-standard registry of people receiving treatment for end-stage kidney disease (the UKRR) enabled us to reduce misclassification of treatment modality (e.g. ∼65% of the kidney transplant subgroup in this study also had a prior dialysis code in their primary care record) and provided a valuable opportunity to audit vaccine implementation and performance within this high-risk population. The integration of UKRR data within OpenSAFELY allows novel linkages to be made with primary care and COVID-19-related outcome data, forming the key foundation for the present analysis.

Given the higher rates of COVID-19-related outcomes following AZ–AZ than BNT–BNT, we cannot rule out the potential influence of selection bias in the three-dose cohort. To mitigate the potential influence of hybrid immunity (which would have been more common in AZ-primed individuals), we opted to exclude individuals if they had any evidence of SARS-CoV-2 infection between doses 1 and 3. The resulting cohort may therefore be unrepresentative of the initial population of AZ–AZ recipients, introducing a bias towards individuals who responded more robustly to the two-dose series or had a lower exposure risk post-vaccination. The phased vaccine roll-out in England also imposed constraints on our analyses. Individuals over 80 years of age, many of whom would have been eligible for this study, were excluded given that BNT had been widely distributed to this age group before AZ became available. Despite careful measures to control for confounding, residual biases may have contributed to the observed discrepancies between vaccine groups. Among people with CKD3, deaths from causes other than COVID-19 were more common in AZ–AZ than BNT–BNT recipients, potentially reflecting greater underlying frailty in this population that was not captured by the clinical covariates (e.g. reflecting easier transport of AZ to individuals unable to attend vaccination centres in person). However, this discrepancy was much smaller than that observed for COVID-19-related outcomes and was not apparent in people with CKD4–5 or in RRT recipients.

Overall, our study serves as an important demonstration of the value of heterologous BNT booster doses (and likely heterologous RNA vaccine booster doses more broadly) in vulnerable populations, suggesting these doses have the capacity to close the gap in immunity observed after prior doses. In settings such as the UK, RNA vaccines have been offered to vulnerable populations across multiple booster campaigns. However, targeted measures to improve the uptake of these doses may be beneficial, particularly in non-White ethnic groups and in deprived settings that have been persistently linked with lower COVID-19 vaccine coverage^29,30^. Our findings also highlight potential ways to optimise vaccination strategies in countries where RNA vaccines make up a portion of the COVID-19 vaccine supply. In particular, administering available RNA vaccines as heterologous second or third doses in high-risk populations might offer greater protection per dose than offering multiple RNA doses to unvaccinated individuals.

## Data Availability

Detailed pseudonymised patient data is potentially re-identifiable and therefore not shared. Access to the underlying identifiable and potentially re-identifiable pseudonymised electronic health record data is tightly governed by various legislative and regulatory frameworks, and restricted by best practice. The data in OpenSAFELY is drawn from General Practice data across England where TPP is the data processor. TPP developers initiate an automated process to create pseudonymised records in the core OpenSAFELY database, which are copies of key structured data tables in the identifiable records. These pseudonymised records are linked onto key external data resources that have also been pseudonymised via SHA-512 one-way hashing of NHS numbers using a shared salt. Bennett Institute for Applied Data Science developers and PIs holding contracts with NHS England have access to the OpenSAFELY pseudonymised data tables as needed to develop the OpenSAFELY tools. These tools in turn enable researchers with OpenSAFELY data access agreements to write and execute code for data management and data analysis without direct access to the underlying raw pseudonymised patient data, and to review the outputs of this code. All code for the full data management pipeline - from raw data to completed results for this analysis - and for the OpenSAFELY platform as a whole is available for review at github.com/OpenSAFELY.

## Acknowledgements

We are very grateful for all the support received from the TPP Technical Operations team throughout this work, and for generous assistance from the information governance and database teams at NHS England/NHS Transformation Directorate.

## Contributors

Conceptualisation: EPKP, FL, SL, DN, and LAT; Methodology: EPKP, EMFH, WJH, JT, BZ, BG, JACS, DN, and LAT; Software: EPKP, EMFH, and WJH; Formal analysis, EPKP and EMFH; Data curation: AM, SS, RS, BG, and DN; Writing–original draft, EPKP; Writing– review & editing, EMFH, WJH, EJC, FL, SL, VM, BM, MS, SS, RS, DN, and LAT; Visualisation, EPKP and WJH; Supervision, DN and LAT; Project administration: EPKP, AM, DN, and LAT; Funding acquisition, EPKP, BG, DN, and LAT.

## Ethical approval

This study was approved by the Health Research Authority (REC reference 20/LO/0651) and by the London School of Hygiene & Tropical Medicine’s Ethics Board (reference 21863).

## Information governance

NHS England is the data controller for OpenSAFELY-TPP; TPP is the data processor; all study authors using OpenSAFELY have the approval of NHS England. This implementation of OpenSAFELY is hosted within the TPP environment which is accredited to the ISO 27001 information security standard and is NHS IG Toolkit compliant. Patient data has been pseudonymised for analysis and linkage using industry standard cryptographic hashing techniques; all pseudonymised datasets transmitted for linkage onto OpenSAFELY are encrypted; access to the platform is via a virtual private network (VPN) connection, restricted to a small group of researchers; the researchers hold contracts with NHS England and only access the platform to initiate database queries and statistical models; all database activity is logged; only aggregate statistical outputs leave the platform environment following best practice for anonymisation of results such as statistical disclosure control for low cell counts.

The OpenSAFELY research platform adheres to the obligations of the UK General Data Protection Regulation (GDPR) and the Data Protection Act 2018. In March 2020, the Secretary of State for Health and Social Care used powers under the UK Health Service (Control of Patient Information) Regulations 2002 (COPI) to require organisations to process confidential patient information for the purposes of protecting public health, providing healthcare services to the public and monitoring and managing the COVID-19 outbreak and incidents of exposure; this sets aside the requirement for patient consent.

Taken together, these provide the legal bases to link patient datasets on the OpenSAFELY platform. GP practices, from which the primary care data are obtained, are required to share relevant health information to support the public health response to the pandemic, and have been informed of the OpenSAFELY analytics platform.

This project includes data from the UKRR derived from patient-level information collected by the NHS as part of the care and support of kidney patients. We thank all kidney patients and kidney centres involved. The data are collated, maintained, and quality assured by the UKRR, which is part of the UK Kidney Association. Access to the data was facilitated by the UKRR’s Data Release Group after consultation with the UK Kidney Association Patient Council. UKRR data are used within OpenSAFELY to address a limited number of critical audit and service delivery questions related to the impact of COVID-19 on patients with kidney disease.

## Data sharing

Detailed pseudonymised patient data is potentially re-identifiable and therefore not shared. Access to the underlying identifiable and potentially re-identifiable pseudonymised electronic health record data is tightly governed by various legislative and regulatory frameworks, and restricted by best practice. The data in OpenSAFELY is drawn from General Practice data across England where TPP is the data processor. TPP developers initiate an automated process to create pseudonymised records in the core OpenSAFELY database, which are copies of key structured data tables in the identifiable records. These pseudonymised records are linked onto key external data resources that have also been pseudonymised via SHA-512 one-way hashing of NHS numbers using a shared salt. Bennett Institute for Applied Data Science developers and PIs holding contracts with NHS England have access to the OpenSAFELY pseudonymised data tables as needed to develop the OpenSAFELY tools. These tools in turn enable researchers with OpenSAFELY data access agreements to write and execute code for data management and data analysis without direct access to the underlying raw pseudonymised patient data, and to review the outputs of this code. All code for the full data management pipeline – from raw data to completed results for this analysis – and for the OpenSAFELY platform as a whole is available for review at github.com/OpenSAFELY.

## Code availability

Data management was performed using Python, with analysis carried out using R 4.0.2. Code for data management and analysis, as well as codelists, are archived online (https://github.com/opensafely/ckd-coverage-ve).

