## Supplementary Materials for "Comparative effectiveness of two- and three-dose schedules involving AZD1222 and BNT162b2 in people with kidney disease: a linked OpenSAFELY and UK Renal Registry cohort study"

**Contents**

Page 2: Figure S1. Cohort characteristics.
Page 3: Figure S2. Cumulative incidence rates by vaccine group in unmatched cohort.
Page 4: Table S1. Definition of key variables.
Page 7: Table S2. Covariate structure of two-dose cohort models.
Page 8: Table S3. Covariate structure of three-dose cohort models.
Page 9: Table S4. Cohort selection.
Page 10: Table S5. Baseline characteristics of three-dose cohort.
Page 11: Table S6. Baseline characteristics of two-dose cohort subgroups.
Page 13: Table S7. Baseline characteristics of three-dose cohort subgroups.
Page 15: Table S8. Incidence rates and hazard ratios for two-dose cohort.
Page 16: Table S9. Incidence rates and hazard ratios for kidney disease subgroups (two-dose cohort).
Page 17: Table S10. Incidence rates and hazard ratios for three-dose cohort.
Page 18: Table S11. Incidence rates and hazard ratios for kidney disease subgroups (three-dose cohort).

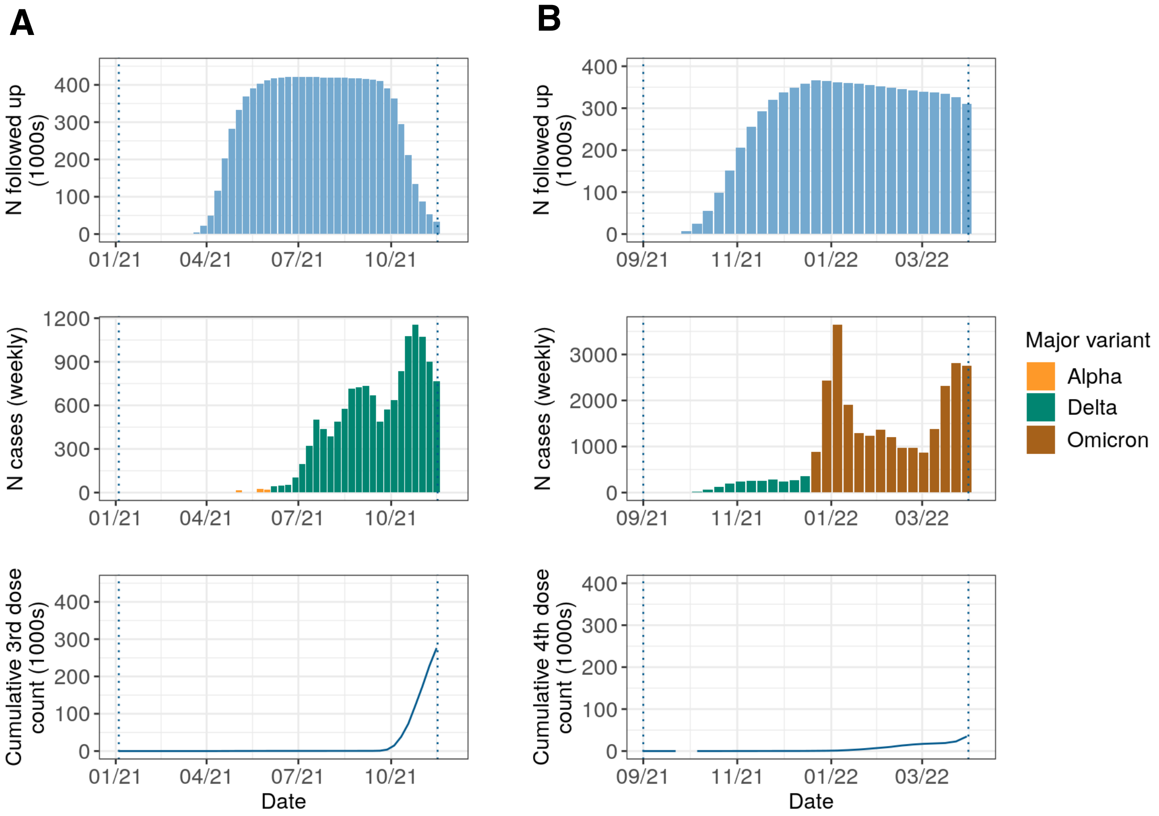


**Figure S1. Cohort characteristics.** Data are shown for (A) two-dose cohort and (B) three-dose cohort. Plots display number of participants enrolled over time (top panels), weekly SARS-CoV-2 infection counts by dominant variant (middle panels), and number of enrolled participants receiving a subsequent dose (bottom panels). Individuals may have experienced outcome events or fulfilled 182 days of follow-up prior to receipt of a subsequent dose. Vertical dotted lines indicate the start of enrolment and end of follow-up for each analysis. Data are rounded to the nearest 5 and non-zero counts of ≤10 redacted. Variant eras were defined as follows: Alpha, up to 3rd June 2021; Delta, 4th June to 14th December 2021; Omicron, 15th December 2021 onwards.

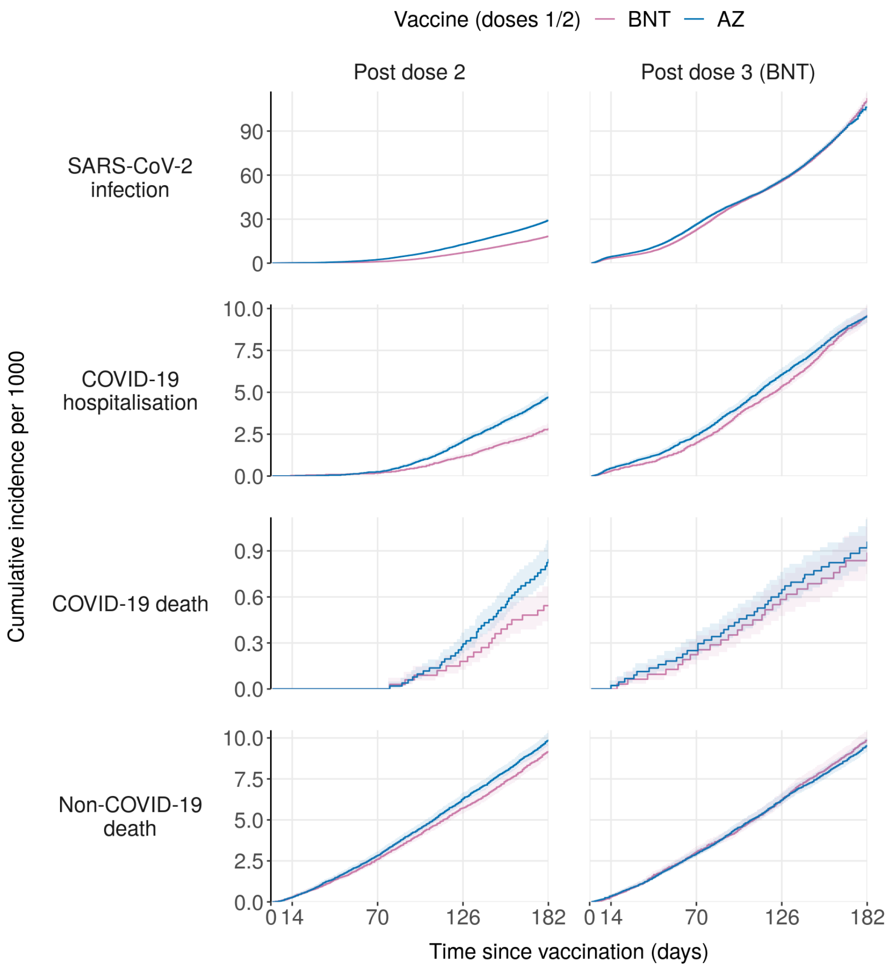


**Figure S2. Cumulative incidence rates by vaccine group in unmatched cohorts.** Kaplan-Meier estimates of cumulative incidence in two-dose and three-dose cohorts are displayed. Kaplan-Meier steps are delayed until ≥5 events occur in compliance with re-identification minimisation requirements in OpenSAFELY. See **Figure 1** for equivalent plots relating to the matched sub-cohorts. Numbers at risk at days 0, 14, 70, and 126 are provided in **Tables S8** (two-dose cohort) and **S10** (three-dose cohort). AZ, AZD1222 (AstraZeneca); BNT, BNT162b2 (Pfizer/BioNTech).

**Table S1. Definition of key variables.**

| Variable | Notes | Values | Date defined (two-dose cohort) | Date defined (three-dose cohort) | Codelist |
|---|---|---|---|---|---|
| **Demography** | | | | | |
| Sex | – | Male; Female | – | – | – |
| Age | Age as integer | Integer (≥16) | 31/03/2021* | 31/03/2021* | – |
| Ethnicity | Derived from primary care record or (if missing) SUS | White; Black; South Asian; Mixed; Other | – | – | opensafely/ethnicity/2020-04-27 |
| NHS region | Derived from practice address | East of England; Midlands; London; North East and Yorkshire; North West; South East; South West | Date of dose 1 | Date of dose 3 | – |
| Index of multiple deprivation | Social deprivation quintile derived from individual's address at Lower Super Output Area (a small geographical area defined by the ONS) | 1 (most deprived); 2; 3; 4; 5 (least deprived) | Date of dose 1 | Date of dose 3 | – |
| Setting (urban/rural) | Derived from individual's address | Urban; Urban conurbation; Rural | Date of dose 1 | Date of dose 3 | – |
| **Exclusion criteria** | | | | | |
| Receipt of end-of-life care | Any prior code corresponding to end-of-life care or midazolam injection (used in end-of-life care) | 0; 1 | Date of dose 1 | Date of dose 3 | opensafely/midazolam-end-of-life/4c1b3c89; nhsd-primary-care-domain-refsets/palcare_cod/5fce98cf |
| Care home residency | Based on matching of individual's address with care homes in CQC database or presence of code in primary record before date defined | 0; 1 | Date of dose 1 | Date of dose 3 | primis-covid19-vacc-uptake/longres/v1 |
| Health or social care worker | Recorded at the time of vaccination | 0; 1 | Any vaccination | Any vaccination | |
| Medically housebound | Any prior code in primary care record with no subsequent code indicating that the individual was no longer housebound | 0; 1 | Date of dose 1 | Date of dose 3 | opensafely/housebound/5bc77310; opensafely/no-longer-housebound/29a88ca6 |
| **Kidney disease status** | | | | | |
| Creatinine level | Serum creatinine level (in μmol/l) as basis for calculation of estimated glomerular filtrate rate (alongside sex and age) | Continuous variable with associated date and operators (>, ~, =) | Last record in 2 years before date of dose 1 | Last record in 2 years before date of dose 3 | CTV3 code XE2q5 |
| Age at creatinine measurement | Age as integer on date of creatinine measurement (used for calculation of estimated glomerular filtrate rate) | Integer | – | – | – |
| UKRR status | Treatment modality in 2020 UKRR prevalence cohort | Dialysis; Transplant | 31/12/2020 | 31/12/2020 | – |
| CKD3-5 (primary care) | Any prior code in primary care record | 0; 1 | Date of dose 1 | Date of dose 3 | primis-covid19-vacc-uptake/ckd35/v1 |
| Dialysis (primary care) | Any prior code in primary care record (individuals with these codes were excluded if absent from UKRR prevalence cohort) | 0; 1 | Date of dose 1 | Date of dose 3 | opensafely/dialysis/3ce108ac |
| Kidney transplant (primary care) | Any prior code in primary care record (individuals with these codes were excluded if absent from UKRR prevalence cohort) | 0; 1 | Date of dose 1 | Date of dose 3 | opensafely/kidney-transplant/2020-07-15 |

| Variable | Notes | Values | Date defined (two-dose cohort) | Date defined (three-dose cohort) | Codelist |
|---|---|---|---|---|---|
| **Clinical comorbidities** | | | | | |
| Immunosuppression | Any prior code indicating the diagnosis of an immunosuppressive condition or any code indicating prescription of immunosuppressive medication in the preceding 6 months | 0; 1 | Date of dose 1 | Date of dose 3 | primis-covid19-vacc-uptake/immdx_cov/v1; primis-covid19-vacc-uptake/immrx/v1 |
| Severe obesity | BMI ≥40 based on most recent weight measurement or clinically coded severe obesity based on most recent BMI-related term in primary care record | 0; 1 | Date of dose 1 | Date of dose 3 | primis-covid19-vacc-uptake/bmi/v1; primis-covid19-vacc-uptake/bmi_stage/v1.2; primis-covid19-vacc-uptake/sev_obesity/v1.2 |
| Diabetes | Any prior code indicating diagnosis of diabetes with no subsequent code indicating that diabetes had resolved | 0; 1 | Date of dose 1 | Date of dose 3 | primis-covid19-vacc-uptake/diab/v1; primis-covid19-vacc-uptake/dmres/v1 |
| Asthma | Any prior emergency admission for asthma or any prior asthma diagnosis in conjunction with asthma systemic steroid prescriptions in each of the preceding 3 months | 0; 1 | Date of dose 1 | Date of dose 3 | primis-covid19-vacc-uptake/ast/v1; primis-covid19-vacc-uptake/astadm/v1; primis-covid19-vacc-uptake/astrx/v1 |
| Chronic respiratory disease | Any prior code in primary care record | 0; 1 | Date of dose 1 | Date of dose 3 | primis-covid19-vacc-uptake/resp_cov/v1 |
| Chronic heart disease | Any prior code in primary care record | 0; 1 | Date of dose 1 | Date of dose 3 | primis-covid19-vacc-uptake/chd_cov/v1.2.1 |
| Chronic liver disease | Any prior code in primary care record | 0; 1 | Date of dose 1 | Date of dose 3 | primis-covid19-vacc-uptake/cld/v1 |
| Asplenia | Any prior code in primary care record | 0; 1 | Date of dose 1 | Date of dose 3 | primis-covid19-vacc-uptake/spln_cov/v1 |
| Haematologic cancer | Any prior code in primary care record | 0; 1 | Date of dose 1 | Date of dose 3 | opensafely/haematological-cancer/2020-04-15 |
| Organ transplant (non-kidney) | Any prior code in primary care record in absence of any indication of kidney transplant (based on primary care code or UK Renal Registry status) | 0; 1 | Date of dose 1 | Date of dose 3 | opensafely/solid-organ-transplantation-snomed/2020-04-10; opensafely/other-organ-transplant/79caeeee |
| Chronic neurological disease including significant learning disorders | Any prior code in primary care record | 0; 1 | Date of dose 1 | Date of dose 3 | primis-covid19-vacc-uptake/cns_cov/v1 |
| Learning disability | Any prior code in primary care record | 0; 1 | Date of dose 1 | Date of dose 3 | primis-covid19-vacc-uptake/learndis/v1 |
| Severe mental illness | Any prior code indicating diagnosis of severe mental illness with no subsequent code indicating that the illness was in remission | 0; 1 | Date of dose 1 | Date of dose 3 | primis-covid19-vacc-uptake/sev_mental/v1; primis-covid19-vacc-uptake/smhres/v1 |
| Clinically extremely vulnerable | Code in primary care record indicating that individual was at high risk of developing complications from COVID-19, with no subsequent code indicating that they were no longer at high risk | 0; 1 | Date of dose 1 | Date of dose 3 | primis-covid19-vacc-uptake/shield/v1; primis-covid19-vacc-uptake/nonshield/v1 |
| **Vaccination status** | COVID-19 vaccination records in NIMS are transferred to an individual's primary care record within days. Sequential vaccination dates were extracted for any COVID-19 vaccine and for specific products (BNT162b2, ChAdOx1-S, and mRNA-1273) | Date + product (Any; BNT162b2; ChAdOx1-S; mRNA-1273) | From 04/01/2021 | From 04/01/2021 | |

| Variable | Notes | Values | Date defined (two-dose cohort) | Date defined (three-dose cohort) | Codelist |
|---|---|---|---|---|---|
| **JCVI group** | Priority groups 1–6 based on criteria defined by the JCVI (groups 1 and 2 were excluded from the analysis) | 1 (care home resident or health/social care worker); 2 (80+); 3 (75+); 4 (70+ or clinically extremely vulnerable); 5 (65+); 6 (16-65 and clinically vulnerable) | Derived from other variables defined above | Derived from other variables defined above | – |
| **Evidence of prior SARS-CoV-2 infection** | Any prior record of a positive SARS-CoV-2 test, COVID-19-related hospitalisation (as defined below), COVID-19-related A&E attendance (defined using the SNOMED code 1240751000000100), or probable COVID-19 (based on primary care coding) | Date | Date of dose 1 | Date of dose 1 (individuals with evidence of SARS-CoV-2 infection between doses 1 and 3 were excluded) | opensafely/covid-identification-in-primary-care-probable-covid-clinical-code/2020-07-16; opensafely/covid-identification-in-primary-care-probable-covid-positive-test/2020-07-16; opensafely/covid-identification-in-primary-care-probable-covid-sequelae/2020-07-16 |
| **No. of SARS-CoV-2 tests** | Number of SGSS records with swab dates in the past 90 dates (including both PCR and lateral flow tests). Fixed date ranges were used in each cohort to avoid the potential influence of changing SARS-CoV-2 incidence on testing patterns. | 0; 1; 2; 3+ | 04/01/2021 | 01/09/2021 | – |
| **Post-vaccination outcomes** | | | | | |
| SARS-CoV-2 positive test | Any positive SARS-CoV-2 test in SGSS based on PCR or lateral flow test. No differentiation is made between asymptomatic and symptomatic infections. Swab date is used as the event date. | Date | From dose 2 | From dose 3 | – |
| COVID-19-related hospital admission | Any completed hospital episode with a COVID-19-related diagnostic code mentioned anywhere in the diagnosis field and with an admission method in the following list: ”21”, ”22”, ”23”, ”24”, ”25”, ”2A”, ”2B”, ”2C”, ”2D”, ”28” | Date | From dose 2 | From dose 3 | opensafely/covid-identification/2020-06-03 |
| COVID-19-related death | Deaths with a COVID-19-related diagnostic code mentioned anywhere on the death certificate (not just as the underlying cause) | Date | From dose 2 | From dose 3 | opensafely/covid-identification/2020-06-03/ |
| Deregistration | Used for censoring | Date | From dose 2 | From dose 3 | – |
| Non-COVID-19 death | Death from any cause as registered in ONS, excluding COVID-19-related deaths as defined above | Date | From dose 2 | From dose 3 | – |

Additional details on variable coding can be found in the full analysis scripts (available at https://github.com/opensafely/ckd-coverage-ve). Codelists can be found at https://codelists.opensafely.org/codelist/[Codelist ID], substituting [Codelist ID] with the identifier listed above.* Age was calculated on 31st March 2021 as per UK Health Security Agency recommendations; for consistency, this was applied to both two- and three-dose cohorts, although priority groups for the autumn 2021 booster campaign were defined based on age as of 31st August 2021. CQC, Care and Quality Commission; JCVI, Joint Committee on Vaccination and Immunisation; ONS, Office for National Statistics; NIMS; National Immunisation Management Service; SGSS, Second Generation Surveillance System; SUS, Secondary Use Service.

**Table S2. Covariate structure of two-dose cohort models.**

| | SARS-CoV-2 infection | | | | | | COVID-19-related hospitalisation | | | | | |
|---|---|---|---|---|---|---|---|---|---|---|---|---|
| | All | CKD3 | CKD4-5 | RRT (transplant) | RRT (dialysis) | RRT (combined) | All | CKD3 | CKD4-5 | RRT (transplant) | RRT (dialysis) | RRT (combined) |
| Vaccination date | Included | Included | Included | Included | Included | Included | Included | Included | Included | Included | Included | Included |
| Age | Included | Included | Included | Included | Included | Included | Included | Included | Included | Included | Included | Included |
| Sex | Included | Included | Included | Included | Included | Included | Included | Included | Included | Included | Included | Included |
| Ethnicity | Included | Included | Included (merged) | Included (merged) | Included (merged) | Included (merged) | Included (merged) | Included (merged) | Included (merged) | Included (merged) | Included (merged) | Included (merged) |
| IMD | Included | Included | Included | Included | Included | Included | Included | Included | Included | Included | Included (merged) | Included |
| Setting (rurality) | Included | Included | Included | Included | Included | Included | Included | Included | Included | Included | Included | Included |
| Kidney disease subgroup | Included | – | – | – | – | – | Included | – | – | – | – | – |
| Immunosuppression | Included | Included | Included | Included | Included | Included | Included | Included | Included | Included | Included | Included |
| Severe obesity | Included | Included | Included | Included | Included | Included | Included | Included | Included | Excluded | Excluded | Excluded |
| Diabetes | Included | Included | Included | Included | Included | Included | Included | Included | Included | Included | Included | Included |
| Chronic respiratory disease (inc. asthma) | Included | Included | Included | Included | Included | Included | Included | Included | Included | Included | Included | Included |
| Chronic heart disease | Included | Included | Included | Included | Included | Included | Included | Included | Included | Included | Included | Included |
| Chronic liver disease | Included | Included | Included | Included | Included | Included | Included | Included | Included | Excluded | Excluded | Included |
| Asplenia | Included | Included | Included | Included | Excluded | Included | Included | Included | Excluded | Excluded | Excluded | Excluded |
| Haematologic cancer | Included | Included | Included | Included | Excluded | Included | Included | Included | Excluded | Excluded | Excluded | Excluded |
| Organ transplant (non-kidney) | Included | Included | Excluded | – | Excluded | Excluded | Included | Included | Excluded | – | Excluded | Excluded |
| Chronic neurological disease | Included | Included | Included | Included | Included | Included | Included | Included | Included | Included | Included | Included |
| Learning disability | Included | Included | Excluded | Included | Excluded | Included | Included | Excluded | Included | Excluded | Excluded | Excluded |
| Severe mental illness | Included | Included | Included | Excluded | Excluded | Excluded | Included | Included | Excluded | Excluded | Excluded | Excluded |
| Prior SARS-CoV-2 | Included | Included | Excluded | Excluded | Excluded | Included | Included | Included | Excluded | Excluded | Excluded | Excluded |
| Number of tests in 90-day window | Included | Included | Included | Included | Included | Included | Included | Included | Included (merged) | Included | Included (merged) | Included |

| | COVID-19-related death | | | | | | Non-COVID-19 death | | | | | |
|---|---|---|---|---|---|---|---|---|---|---|---|---|
| | All | CKD3 | CKD4-5 | RRT (transplant) | RRT (dialysis) | RRT (combined) | All | CKD3 | CKD4-5 | RRT (transplant) | RRT (dialysis) | RRT (combined) |
| Vaccination date | Included | Included | [R] | Included | [R] | Included | Included | Included | Included | Included | Included | Included |
| Age | Included | Included | [R] | Included | [R] | Included | Included | Included | Included | Included | Included | Included |
| Sex | Included | Included | [R] | Included | [R] | Included | Included | Included | Included | Included | Included | Included |
| Ethnicity | Included (merged) | Included (merged) | [R] | Included (merged) | [R] | Included (merged) | Included | Included | Included (merged) | Included (merged) | Included (merged) | Included (merged) |
| IMD | Included | Included | [R] | Included (merged) | [R] | Included | Included | Included | Included | Included | Included | Included |
| Setting (rurality) | Included | Included | [R] | Included | [R] | Included | Included | Included | Included | Included | Included | Included |
| Kidney disease subgroup | Included | – | [R] | – | [R] | – | Included | – | – | – | – | – |
| Immunosuppression | Included | Included | [R] | Included | [R] | Included | Included | Included | Included | Included | Included | Included |
| Severe obesity | Included | Included | [R] | Excluded | [R] | Excluded | Included | Included | Included | Excluded | Included | Included |
| Diabetes | Included | Included | [R] | Included | [R] | Included | Included | Included | Included | Included | Included | Included |
| Chronic respiratory disease (inc. asthma) | Included | Included | [R] | Included | [R] | Included | Included | Included | Included | Included | Included | Included |
| Chronic heart disease | Included | Included | [R] | Excluded | [R] | Excluded | Included | Included | Included | Included | Included | Included |
| Chronic liver disease | Included | Excluded | [R] | Excluded | [R] | Excluded | Included | Included | Included | Included | Included | Included |
| Asplenia | Excluded | Excluded | [R] | Excluded | [R] | Excluded | Included | Included | Excluded | Excluded | Excluded | Included |
| Haematologic cancer | Included | Included | [R] | Excluded | [R] | Excluded | Included | Included | Included | Excluded | Included | Included |
| Organ transplant (non-kidney) | Excluded | Excluded | [R] | Excluded | [R] | Excluded | Included | Included | Excluded | – | Excluded | Excluded |
| Chronic neurological disease | Included | Included | [R] | Excluded | [R] | Excluded | Included | Included | Included | Included | Included | Included |
| Learning disability | Excluded | Excluded | [R] | Excluded | [R] | Excluded | Included | Included | Excluded | Excluded | Excluded | Excluded |
| Severe mental illness | Excluded | Excluded | [R] | Excluded | [R] | Excluded | Included | Included | Excluded | Excluded | Excluded | Excluded |
| Prior SARS-CoV-2 | Excluded | Excluded | [R] | Excluded | [R] | Excluded | Included | Included | Included | Excluded | Included | Included |
| Number of tests in 90-day window | Included (merged) | Included (merged) | [R] | Included (merged) | [R] | Included (merged) | Included | Included | Included | Included (merged) | Included | Included |

We excluded binary covariates if cross-tabulating the variable with vaccine group yielded any cell with fewer than three events. For categorical variables with more than two levels, we merged categories until all levels fulfilled the cross-tabulation criteria described above or only one level remained, in which case the variable was excluded. CKD, chronic kidney disease; IMD, index of multiple deprivation; [R], redacted due to low event counts in one or both vaccine groups; RRT, renal replacement therapy.

**Key**

| | |
|---|---|
| (green) | Included |
| (blue) | Included with one or more covariate levels merged |
| (red) | Excluded due to insufficient event counts across covariate levels |
| – | Excluded by design |

**Table S3. Covariate structure of three-dose cohort models.**

| | SARS-CoV-2 infection | | | | | | COVID-19-related hospitalisation | | | | | | COVID-19-related death | | | | | | Non-COVID-19 death | | | | | |
|---|---|---|---|---|---|---|---|---|---|---|---|---|---|---|---|---|---|---|---|---|---|---|---|---|
| | All | CKD3 | CKD4-5 | RRT (transplant) | RRT (dialysis) | RRT (combined) | All | CKD3 | CKD4-5 | RRT (transplant) | RRT (dialysis) | RRT (combined) | All | CKD3 | CKD4-5 | RRT (transplant) | RRT (dialysis) | RRT (combined) | All | CKD3 | CKD4-5 | RRT (transplant) | RRT (dialysis) | RRT (combined) |
| Vaccination date | Included | Included | Included | Included | Included | Included | Included | Included | Included | Included | Included | Included | Included | Included | Included | Included | | Included | Included | Included | Included | Included | Included | Included |
| Age | Included | Included | Included | Included | Included | Included | Included | Included | Included | Included | Included | Included | Included | Included | Included | Included | | Included | Included | Included | Included | Included | Included | Included |
| Sex | Included | Included | Included | Included | Included | Included | Included | Included | Included | Included | Included | Included | Included | Included | Included | Included | | Included | Included | Included | Included | Included | Included | Included |
| Ethnicity | Included | Included | Included | Included | Included | Included | Included | Merged | Merged | Merged | Merged | Merged | Merged | Merged | Excluded | Merged | | Merged | Included | Merged | Merged | Merged | Merged | Merged |
| IMD | Included | Included | Included | Included | Included | Included | Included | Included | Included | Included | Included | Included | Included | Included | Merged | Merged | | Merged | Included | Included | Included | Included | Included | Included |
| Setting (rurality) | Included | Included | Included | Included | Included | Included | Included | Included | Included | Included | Included | Included | Included | Included | Included | Merged | | Merged | Included | Included | Included | Included | Included | Included |
| Kidney disease subgroup | Included | – | – | – | – | – | Included | – | – | – | – | – | Included | Excluded | Excluded | Excluded | | Excluded | Included | – | – | – | – | – |
| Immunosuppression | Included | Included | Included | Included | Included | Included | Included | Included | Included | Included | Included | Included | Included | Included | Excluded | Included | | Included | Included | Included | Included | Included | Included | Included |
| Severe obesity | Included | Included | Included | Included | Included | Included | Included | Included | Included | Included | Excluded | Included | Included | Included | Excluded | Excluded | | Excluded | Included | Included | Included | Excluded | Included | Included |
| Diabetes | Included | Included | Included | Included | Included | Included | Included | Included | Included | Included | Included | Included | Included | Included | Included | Included | | Included | Included | Included | Included | Included | Included | Included |
| Chronic respiratory disease (inc. asthma) | Included | Included | Included | Included | Included | Included | Included | Included | Included | Included | Included | Included | Included | Included | Included | Included | [R] | Excluded | Included | Included | Included | Included | Included | Included |
| Chronic heart disease | Included | Included | Included | Included | Included | Included | Included | Included | Included | Included | Included | Included | Included | Included | Included | Included | | Included | Included | Included | Included | Included | Included | Included |
| Chronic liver disease | Included | Included | Included | Included | Included | Included | Included | Included | Included | Included | Excluded | Included | Included | Included | Excluded | Excluded | | Excluded | Included | Included | Included | Excluded | Included | Included |
| Asplenia | Included | Included | Included | Included | Included | Included | Included | Included | Excluded | Included | Excluded | Excluded | Excluded | Excluded | Included | Included | | Excluded | Included | Included | Excluded | Excluded | Excluded | Excluded |
| Haematologic cancer | Included | Included | Included | Included | Included | Included | Included | Included | Included | Excluded | Included | Included | Included | Included | Excluded | Excluded | | Excluded | Included | Included | Included | Excluded | Excluded | Included |
| Organ transplant (non-kidney) | Included | Included | Included | – | Excluded | Excluded | Included | Included | Included | – | Excluded | Excluded | Excluded | Excluded | Excluded | Excluded | | Excluded | Included | Included | Excluded | – | Excluded | Excluded |
| Chronic neurological disease | Included | Included | Included | Included | Included | Included | Included | Included | Included | Included | Included | Included | Included | Included | Excluded | Excluded | | Included | Included | Included | Included | Included | Included | Included |
| Learning disability | Included | Included | Excluded | Included | Included | Included | Included | Included | Excluded | Excluded | Excluded | Excluded | Excluded | Excluded | Excluded | Excluded | | Excluded | Included | Included | Excluded | Excluded | Excluded | Excluded |
| Severe mental illness | Included | Included | Included | Excluded | Included | Included | Included | Included | Excluded | Excluded | Excluded | Excluded | Excluded | Excluded | Excluded | Excluded | | Excluded | Included | Included | Excluded | Excluded | Excluded | Excluded |
| Prior SARS-CoV-2 | Included | Included | Included | Included | Included | Included | Included | Included | Included | Included | Included | Included | Excluded | Excluded | Excluded | Excluded | | Excluded | Included | Included | Included | Included | Included | Included |
| Number of tests in 90-day window | Included | Included | Included | Included | Included | Included | Included | Included | Included | Included | Included | Included | Included | Included | Excluded | Merged | | Merged | Included | Included | Included | Merged | Merged | Included |

We excluded binary covariates if cross-tabulating the variable with vaccine group yielded any cell with fewer than three events. For categorical variables with more than two levels, we merged categories until all levels fulfilled the cross-tabulation criteria described above or only one level remained, in which case the variable was excluded. CKD, chronic kidney disease; IMD, index of multiple deprivation; [R], redacted due to low event counts in one or both vaccine groups; RRT, renal replacement therapy.

**Key**

| | |
|---|---|
| | Included |
| | Included with one or more covariate levels merged |
| | Excluded due to insufficient event counts across covariate levels |
| – | Excluded by design |

**Table S4. Cohort selection.**

| Criteria | N retained, two-dose cohort | N retained, three-dose cohort |
|---|---|---|
| (Registered for at least 3 months prior to first COVID-19 vaccine dose *AND* aged ≥16 years on 31st March 2021)<br>*AND*<br>(Received first dose on or after 4th January 2021 [two-dose cohort] or third dose on or after 1st September 2021 [three-dose cohort])<br>*AND*<br>(At least one serum creatinine measurement in 2 years preceding definition date *OR* in UK Renal Registry on 31st December 2020) | 8,301,730 | 7,101,160 |
| Valid record for most recent creatinine measurement (with associated date and no linked operators) *OR* in UK Renal Registry on 31st December 2020 | 8,301,515 | 7,100,995 |
| eGFR <60 ml/min/1.73 $m^2$ based on most recent creatinine measurement *OR* in UK Renal Registry on 31st December 2020 | 1,007,300 | 954,640 |
| No RRT status mismatch (primary care code indicating dialysis or kidney transplant but not in UK Renal Registry on 31st December 2020) | 1,004,705 | 951,300 |
| No missing demographic information (sex, region, index of multiple deprivation, or ethnicity) | 965,555 | 914,280 |
| Received AZ–AZ/BNT–BNT (two-dose cohort) or AZ–AZ–BNT/BNT–BNT–BNT (three-dose cohort) | 919,540 | 834,195 |
| Received second dose on or before 17th October 2021 (two-dose cohort) | 899,410 | – |
| Received first dose on or after 4th January 2021 and third dose between 1st September 2021 and 1st March 2022 (three-dose cohort) | – | 721,285 |
| Dose 1–2 interval of 8–14 weeks (both cohorts) and dose 2–3 interval of ≥12 weeks (three-dose cohort) | 777,810 | 693,575 |
| Not healthcare worker, care home resident, receiving end-of-life care, housebound, or in JCVI priority group 2 | 438,485 | 390,280 |
| No censoring events before start of follow-up | 438,475 | 390,280 |
| No documented SARS-CoV-2 infection in window spanning 90 days before dose 1 (two-dose cohort) | 428,670 | – |
| No documented SARS-CoV-2 infection between doses 1 and 2 (two-dose cohort) or between dose 1 and dose 3 (three-dose cohort) | 426,780 | 377,395 |

Frequencies are rounded to the nearest 5. AZ, AZD1222 (AstraZeneca); BNT, BNT162b2 (Pfizer/BioNTech); CKD, chronic kidney disease; eGFR, estimated glomerular filtrate rate; JCVI, Joint Committee on Vaccination and Immunisation; RRT, renal replacement therapy.

**Table S5. Baseline characteristics of three-dose cohort.**

| Characteristic | Unmatched | | Matched | |
|---|---|---|---|---|
| | **AZ–AZ–BNT N = 220,330** | **BNT–BNT–BNT N = 157,065** | **AZ–AZ–BNT N = 127,345** | **BNT–BNT–BNT N = 127,345** |
| **Age** | | | | |
| 16–64 | 38,255 (17.4%) | 23,345 (14.9%) | 12,415 (9.7%) | 13,625 (10.7%) |
| 65–69 | 34,100 (15.5%) | 19,335 (12.3%) | 16,810 (13.2%) | 16,505 (13.0%) |
| 70–74 | 72,730 (33.0%) | 43,200 (27.5%) | 44,855 (35.2%) | 38,050 (29.9%) |
| 75–79 | 75,245 (34.2%) | 71,190 (45.3%) | 53,265 (41.8%) | 59,165 (46.5%) |
| **Sex** | | | | |
| Female | 115,590 (52.5%) | 82,525 (52.5%) | 67,490 (53.0%) | 67,490 (53.0%) |
| Male | 104,740 (47.5%) | 74,540 (47.5%) | 59,855 (47.0%) | 59,855 (47.0%) |
| **Ethnicity** | | | | |
| White | 207,500 (94.2%) | 148,125 (94.3%) | 121,935 (95.8%) | 122,110 (95.9%) |
| Black | 3,220 (1.5%) | 2,100 (1.3%) | 1,095 (0.9%) | 1,175 (0.9%) |
| South Asian | 7,190 (3.3%) | 5,145 (3.3%) | 3,220 (2.5%) | 2,980 (2.3%) |
| Mixed | 975 (0.4%) | 725 (0.5%) | 405 (0.3%) | 470 (0.4%) |
| Other | 1,450 (0.7%) | 965 (0.6%) | 685 (0.5%) | 610 (0.5%) |
| **Index of multiple deprivation quintile** | | | | |
| 1 most deprived | 35,750 (16.2%) | 24,935 (15.9%) | 19,040 (15.0%) | 19,040 (15.0%) |
| 2 | 41,825 (19.0%) | 29,815 (19.0%) | 23,285 (18.3%) | 23,285 (18.3%) |
| 3 | 50,375 (22.9%) | 36,050 (23.0%) | 29,660 (23.3%) | 29,660 (23.3%) |
| 4 | 48,270 (21.9%) | 34,630 (22.0%) | 28,660 (22.5%) | 28,660 (22.5%) |
| 5 least deprived | 44,105 (20.0%) | 31,635 (20.1%) | 26,700 (21.0%) | 26,700 (21.0%) |
| **Setting** | | | | |
| Urban city or town | 117,970 (53.5%) | 82,915 (52.8%) | 69,570 (54.6%) | 67,620 (53.1%) |
| Urban conurbation | 44,175 (20.0%) | 31,675 (20.2%) | 23,585 (18.5%) | 23,505 (18.5%) |
| Rural | 58,185 (26.4%) | 42,475 (27.0%) | 34,190 (26.8%) | 36,220 (28.4%) |
| **Kidney disease** | | | | |
| CKD3a | 164,575 (74.7%) | 115,730 (73.7%) | 99,715 (78.3%) | 99,715 (78.3%) |
| CKD3b | 39,920 (18.1%) | 30,085 (19.2%) | 22,720 (17.8%) | 22,720 (17.8%) |
| CKD4–5 | 9,605 (4.4%) | 6,885 (4.4%) | 3,555 (2.8%) | 3,555 (2.8%) |
| RRT (dialysis) | 1,785 (0.8%) | 1,480 (0.9%) | 220 (0.2%) | 220 (0.2%) |
| RRT (transplant) | 4,445 (2.0%) | 2,885 (1.8%) | 1,130 (0.9%) | 1,130 (0.9%) |
| **Primary care coding of kidney disease** | | | | |
| CKD3–5 | 121,440 (55.1%) | 90,165 (57.4%) | 70,520 (55.4%) | 71,625 (56.2%) |
| Dialysis code | 4,385 (2.0%) | 3,070 (2.0%) | 900 (0.7%) | 930 (0.7%) |
| Kidney transplant code | 4,715 (2.1%) | 3,100 (2.0%) | 1,115 (0.9%) | 1,125 (0.9%) |
| **Morbidities** | | | | |
| Immunosuppression | 14,610 (6.6%) | 10,245 (6.5%) | 5,435 (4.3%) | 5,455 (4.3%) |
| Severe obesity | 13,185 (6.0%) | 9,360 (6.0%) | 7,040 (5.5%) | 7,210 (5.7%) |
| Diabetes | 62,455 (28.3%) | 45,670 (29.1%) | 35,925 (28.2%) | 35,625 (28.0%) |
| Chronic respiratory disease (inc. asthma) | 25,020 (11.4%) | 18,000 (11.5%) | 14,455 (11.4%) | 14,100 (11.1%) |
| Chronic heart disease | 80,525 (36.5%) | 60,000 (38.2%) | 48,175 (37.8%) | 48,535 (38.1%) |
| Chronic liver disease | 10,130 (4.6%) | 7,065 (4.5%) | 5,470 (4.3%) | 5,410 (4.2%) |
| Asplenia | 2,200 (1.0%) | 1,510 (1.0%) | 860 (0.7%) | 825 (0.6%) |
| Haematologic cancer | 4,535 (2.1%) | 3,415 (2.2%) | 1,890 (1.5%) | 1,900 (1.5%) |
| Organ transplant (non-kidney) | 885 (0.4%) | 460 (0.3%) | 220 (0.2%) | 180 (0.1%) |
| Chronic neurological disease | 23,880 (10.8%) | 17,415 (11.1%) | 13,995 (11.0%) | 13,970 (11.0%) |
| Learning disability | 885 (0.4%) | 485 (0.3%) | 300 (0.2%) | 275 (0.2%) |
| Severe mental illness | 3,015 (1.4%) | 1,850 (1.2%) | 1,435 (1.1%) | 1,420 (1.1%) |
| Clinically extremely vulnerable | 49,575 (22.5%) | 35,755 (22.8%) | 23,010 (18.1%) | 23,010 (18.1%) |
| **Prior SARS-CoV-2 infection** | 8,215 (3.7%) | 4,470 (2.8%) | 1,590 (1.2%) | 1,590 (1.2%) |
| **No. of SARS-CoV-2 tests in 90-day pre-vaccination window** | | | | |
| 0 | 170,755 (77.5%) | 121,775 (77.5%) | 99,830 (78.4%) | 100,750 (79.1%) |
| 1 | 22,520 (10.2%) | 15,985 (10.2%) | 13,070 (10.3%) | 12,675 (10.0%) |
| 2 | 8,640 (3.9%) | 6,000 (3.8%) | 4,915 (3.9%) | 4,665 (3.7%) |
| 3+ | 18,415 (8.4%) | 13,305 (8.5%) | 9,530 (7.5%) | 9,255 (7.3%) |
| **Region** | | | | |
| East of England | 46,510 (21.1%) | 35,880 (22.8%) | 29,125 (22.9%) | 29,125 (22.9%) |
| Midlands | 52,090 (23.6%) | 36,640 (23.3%) | 31,190 (24.5%) | 31,190 (24.5%) |
| London | 4,595 (2.1%) | 5,510 (3.5%) | 2,245 (1.8%) | 2,245 (1.8%) |
| North East and Yorkshire | 42,855 (19.5%) | 28,550 (18.2%) | 23,740 (18.6%) | 23,740 (18.6%) |
| North West | 21,125 (9.6%) | 13,610 (8.7%) | 10,935 (8.6%) | 10,935 (8.6%) |
| South East | 13,280 (6.0%) | 9,005 (5.7%) | 6,575 (5.2%) | 6,575 (5.2%) |
| South West | 39,865 (18.1%) | 27,870 (17.7%) | 23,535 (18.5%) | 23,535 (18.5%) |
| **JCVI priority group** | | | | |
| 3 (75+) | 75,245 (34.2%) | 71,190 (45.3%) | 53,265 (41.8%) | 59,165 (46.5%) |
| 4 (70+ or clinically extremely vulnerable) | 92,995 (42.2%) | 55,325 (35.2%) | 50,815 (39.9%) | 44,005 (34.6%) |
| 5 (65+) | 26,545 (12.0%) | 15,135 (9.6%) | 13,945 (11.0%) | 13,725 (10.8%) |
| 6 (16–65 and clinically vulnerable) | 25,545 (11.6%) | 15,420 (9.8%) | 9,320 (7.3%) | 10,450 (8.2%) |

See **Table S1** for further details on variable definitions. Data are n (%) after rounding to the nearest 5. AZ, AZD1222 (AstraZeneca); BNT, BNT162b2 (Pfizer/BioNTech); CKD, chronic kidney disease; JCVI, Joint Committee on Vaccination and Immunisation; RRT, renal replacement therapy.

**Table S6. Baseline characteristics of two-dose cohort subgroups.**

| | CKD3 | | CKD4–5 | | RRT (transplant) | | RRT (dialysis) | | RRT (combined) | |
|---|---|---|---|---|---|---|---|---|---|---|
| **Characteristic** | **AZ–AZ N = 239,380** | **BNT–BNT N = 156,965** | **AZ–AZ N = 10,325** | **BNT–BNT N = 6,985** | **AZ–AZ N = 5,540** | **BNT–BNT N = 3,385** | **AZ–AZ N = 2,335** | **BNT–BNT N = 1,870** | **AZ–AZ N = 7,875** | **BNT–BNT N = 5,255** |
| **Age** | | | | | | | | | | |
| 16–64 | 46,390 (19.4%) | 24,570 (15.7%) | 2,570 (24.9%) | 1,565 (22.4%) | 4,120 (74.4%) | 2,410 (71.2%) | 1,250 (53.5%) | 940 (50.3%) | 5,370 (68.2%) | 3,350 (63.7%) |
| 65–69 | 39,385 (16.5%) | 20,280 (12.9%) | 1,465 (14.2%) | 825 (11.8%) | 680 (12.3%) | 390 (11.5%) | 360 (15.4%) | 245 (13.1%) | 1,040 (13.2%) | 635 (12.1%) |
| 70–74 | 76,415 (31.9%) | 42,825 (27.3%) | 2,905 (28.1%) | 1,630 (23.3%) | 520 (9.4%) | 365 (10.8%) | 370 (15.8%) | 305 (16.3%) | 890 (11.3%) | 670 (12.7%) |
| 75–79 | 77,190 (32.2%) | 69,290 (44.1%) | 3,380 (32.7%) | 2,965 (42.4%) | 225 (4.1%) | 220 (6.5%) | 355 (15.2%) | 380 (20.3%) | 575 (7.3%) | 600 (11.4%) |
| **Sex** | | | | | | | | | | |
| Female | 128,710 (53.8%) | 84,120 (53.6%) | 4,650 (45.0%) | 3,030 (43.4%) | 2,060 (37.2%) | 1,275 (37.7%) | 880 (37.7%) | 670 (35.8%) | 2,940 (37.3%) | 1,945 (37.0%) |
| Male | 110,665 (46.2%) | 72,845 (46.4%) | 5,675 (55.0%) | 3,955 (56.6%) | 3,485 (62.9%) | 2,110 (62.3%) | 1,455 (62.3%) | 1,200 (64.2%) | 4,940 (62.7%) | 3,310 (63.0%) |
| **Ethnicity** | | | | | | | | | | |
| White | 225,190 (94.1%) | 148,410 (94.5%) | 9,200 (89.1%) | 6,285 (90.0%) | 4,685 (84.6%) | 2,810 (83.0%) | 1,825 (78.2%) | 1,555 (83.2%) | 6,510 (82.7%) | 4,365 (83.1%) |
| Black | 4,145 (1.7%) | 2,210 (1.4%) | 235 (2.3%) | 145 (2.1%) | 160 (2.9%) | 105 (3.1%) | 140 (6.0%) | 75 (4.0%) | 300 (3.8%) | 180 (3.4%) |
| South Asian | 7,265 (3.0%) | 4,670 (3.0%) | 710 (6.9%) | 450 (6.4%) | 545 (9.8%) | 370 (10.9%) | 300 (12.8%) | 195 (10.4%) | 845 (10.7%) | 565 (10.8%) |
| Mixed | 1,220 (0.5%) | 750 (0.5%) | 70 (0.7%) | 45 (0.6%) | 50 (0.9%) | 20 (0.6%) | 25 (1.1%) | 20 (1.1%) | 80 (1.0%) | 40 (0.8%) |
| Other | 1,560 (0.7%) | 925 (0.6%) | 105 (1.0%) | 65 (0.9%) | 100 (1.8%) | 75 (2.2%) | 45 (1.9%) | 30 (1.6%) | 145 (1.8%) | 105 (2.0%) |
| **Index of multiple deprivation quintile** | | | | | | | | | | |
| 1 most deprived | 39,935 (16.7%) | 25,155 (16.0%) | 2,165 (21.0%) | 1,430 (20.5%) | 1,065 (19.2%) | 620 (18.3%) | 690 (29.6%) | 450 (24.1%) | 1,755 (22.3%) | 1,065 (20.3%) |
| 2 | 45,815 (19.1%) | 29,985 (19.1%) | 2,125 (20.6%) | 1,480 (21.2%) | 1,105 (19.9%) | 690 (20.4%) | 545 (23.3%) | 430 (23.0%) | 1,650 (21.0%) | 1,120 (21.3%) |
| 3 | 54,315 (22.7%) | 36,115 (23.0%) | 2,320 (22.5%) | 1,565 (22.4%) | 1,200 (21.7%) | 780 (23.0%) | 460 (19.7%) | 395 (21.1%) | 1,660 (21.1%) | 1,180 (22.5%) |
| 4 | 52,155 (21.8%) | 34,605 (22.0%) | 1,990 (19.3%) | 1,380 (19.8%) | 1,155 (20.8%) | 670 (19.8%) | 355 (15.2%) | 325 (17.4%) | 1,510 (19.2%) | 1,000 (19.0%) |
| 5 least deprived | 47,160 (19.7%) | 31,105 (19.8%) | 1,715 (16.6%) | 1,125 (16.1%) | 1,015 (18.3%) | 620 (18.3%) | 285 (12.2%) | 270 (14.4%) | 1,300 (16.5%) | 890 (16.9%) |
| **Setting** | | | | | | | | | | |
| Urban city or town | 128,970 (53.9%) | 83,545 (53.2%) | 5,390 (52.2%) | 3,615 (51.8%) | 3,015 (54.4%) | 1,815 (53.6%) | 1,120 (48.0%) | 1,080 (57.8%) | 4,135 (52.5%) | 2,895 (55.1%) |
| Urban conurbation | 45,735 (19.1%) | 30,170 (19.2%) | 2,500 (24.2%) | 1,650 (23.6%) | 1,320 (23.8%) | 805 (23.8%) | 790 (33.8%) | 410 (21.9%) | 2,110 (26.8%) | 1,215 (23.1%) |
| Rural | 64,675 (27.0%) | 43,245 (27.6%) | 2,430 (23.5%) | 1,720 (24.6%) | 1,205 (21.8%) | 760 (22.5%) | 425 (18.2%) | 385 (20.6%) | 1,630 (20.7%) | 1,145 (21.8%) |
| **Kidney disease** | | | | | | | | | | |
| CKD3a | 195,125 (81.5%) | 126,335 (80.5%) | [N] | [N] | [N] | [N] | [N] | [N] | [N] | [N] |
| CKD3b | 44,255 (18.5%) | 30,630 (19.5%) | [N] | [N] | [N] | [N] | [N] | [N] | [N] | [N] |
| CKD4–5 | [N] | [N] | 10,325 (100.0%) | 6,985 (100.0%) | [N] | [N] | [N] | [N] | [N] | [N] |
| RRT (dialysis) | [N] | [N] | [N] | [N] | [N] | [N] | 2,335 (100.0%) | 1,870 (100.0%) | 2,335 (29.7%) | 1,870 (35.6%) |
| RRT (transplant) | [N] | [N] | [N] | [N] | 5,540 (100.0%) | 3,385 (100.0%) | [N] | [N] | 5,540 (70.3%) | 3,385 (64.4%) |
| **Primary care coding of kidney disease** | | | | | | | | | | |
| CKD3–5 | 122,315 (51.1%) | 85,685 (54.6%) | 8,945 (86.6%) | 6,150 (88.0%) | 4,170 (75.3%) | 2,625 (77.5%) | 1,965 (84.2%) | 1,565 (83.7%) | 6,135 (77.9%) | 4,190 (79.7%) |
| Dialysis code | [N] | [N] | [N] | [N] | 3,585 (64.7%) | 2,150 (63.5%) | 1,885 (80.7%) | 1,505 (80.5%) | 5,470 (69.5%) | 3,655 (69.6%) |
| Kidney transplant code | [N] | [N] | [N] | [N] | 5,400 (97.5%) | 3,285 (97.0%) | 400 (17.1%) | 300 (16.0%) | 5,800 (73.7%) | 3,585 (68.2%) |
| **Morbidities** | | | | | | | | | | |
| Immunosuppression | 12,980 (5.4%) | 8,820 (5.6%) | 745 (7.2%) | 510 (7.3%) | 2,185 (39.4%) | 1,350 (39.9%) | 245 (10.5%) | 205 (11.0%) | 2,430 (30.9%) | 1,560 (29.7%) |
| Severe obesity | 14,550 (6.1%) | 9,355 (6.0%) | 950 (9.2%) | 600 (8.6%) | 130 (2.3%) | 80 (2.4%) | 185 (7.9%) | 130 (7.0%) | 315 (4.0%) | 210 (4.0%) |
| Diabetes | 63,090 (26.4%) | 42,780 (27.3%) | 4,955 (48.0%) | 3,420 (49.0%) | 1,655 (29.9%) | 1,025 (30.3%) | 990 (42.4%) | 755 (40.4%) | 2,645 (33.6%) | 1,775 (33.8%) |
| Chronic respiratory disease (inc. asthma) | 25,615 (10.7%) | 17,190 (11.0%) | 1,305 (12.6%) | 915 (13.1%) | 410 (7.4%) | 265 (7.8%) | 240 (10.3%) | 215 (11.5%) | 645 (8.2%) | 480 (9.1%) |
| Chronic heart disease | 81,075 (33.9%) | 56,605 (36.1%) | 4,590 (44.5%) | 3,135 (44.9%) | 1,605 (29.0%) | 1,070 (31.6%) | 1,105 (47.3%) | 920 (49.2%) | 2,710 (34.4%) | 1,990 (37.9%) |
| Chronic liver disease | 10,190 (4.3%) | 6,510 (4.1%) | 575 (5.6%) | 375 (5.4%) | 255 (4.6%) | 170 (5.0%) | 145 (6.2%) | 105 (5.6%) | 400 (5.1%) | 275 (5.2%) |
| Asplenia | 2,240 (0.9%) | 1,445 (0.9%) | 130 (1.3%) | 65 (0.9%) | 80 (1.4%) | 45 (1.3%) | 40 (1.7%) | 20 (1.1%) | 120 (1.5%) | 65 (1.2%) |
| Haematologic cancer | 4,230 (1.8%) | 3,010 (1.9%) | 295 (2.9%) | 200 (2.9%) | 85 (1.5%) | 80 (2.4%) | 65 (2.8%) | 65 (3.5%) | 150 (1.9%) | 145 (2.8%) |
| Organ transplant (non-kidney) | 850 (0.4%) | 460 (0.3%) | 95 (0.9%) | 50 (0.7%) | [N] | [N] | 15 (0.6%) | 15 (0.8%) | 15 (0.2%) | 15 (0.3%) |
| Chronic neurological disease | 24,160 (10.1%) | 16,435 (10.5%) | 1,370 (13.3%) | 920 (13.2%) | 510 (9.2%) | 305 (9.0%) | 295 (12.6%) | 260 (13.9%) | 805 (10.2%) | 565 (10.8%) |
| Learning disability | 970 (0.4%) | 495 (0.3%) | 80 (0.8%) | 35 (0.5%) | 65 (1.2%) | 55 (1.6%) | 25 (1.1%) | 15 (0.8%) | 90 (1.1%) | 75 (1.4%) |

| | | | | | | | | | | |
|---|---|---|---|---|---|---|---|---|---|---|
| Severe mental illness | 3,435 (1.4%) | 1,965 (1.3%) | 245 (2.4%) | 120 (1.7%) | 50 (0.9%) | 25 (0.7%) | 55 (2.4%) | 30 (1.6%) | 105 (1.3%) | 55 (1.0%) |
| Clinically extremely vulnerable | 37,170 (15.5%) | 24,900 (15.9%) | 3,815 (36.9%) | 2,435 (34.9%) | 5,480 (98.9%) | 3,345 (98.8%) | 2,270 (97.2%) | 1,795 (96.0%) | 7,750 (98.4%) | 5,140 (97.8%) |
| **Prior SARS-CoV-2 infection** | 3,265 (1.4%) | 1,865 (1.2%) | 245 (2.4%) | 135 (1.9%) | 165 (3.0%) | 105 (3.1%) | 200 (8.6%) | 130 (7.0%) | 365 (4.6%) | 240 (4.6%) |
| **No. of SARS-CoV-2 tests in 90-day pre-vaccination window** | | | | | | | | | | |
| 0 | 208,390 (87.1%) | 136,865 (87.2%) | 8,380 (81.2%) | 5,700 (81.6%) | 4,350 (78.5%) | 2,600 (76.8%) | 825 (35.3%) | 650 (34.8%) | 5,170 (65.7%) | 3,250 (61.8%) |
| 1 | 20,815 (8.7%) | 13,585 (8.7%) | 1,050 (10.2%) | 735 (10.5%) | 695 (12.5%) | 450 (13.3%) | 350 (15.0%) | 250 (13.4%) | 1,045 (13.3%) | 700 (13.3%) |
| 2 | 5,295 (2.2%) | 3,480 (2.2%) | 400 (3.9%) | 265 (3.8%) | 225 (4.1%) | 140 (4.1%) | 175 (7.5%) | 125 (6.7%) | 400 (5.1%) | 265 (5.0%) |
| 3+ | 4,875 (2.0%) | 3,040 (1.9%) | 495 (4.8%) | 285 (4.1%) | 275 (5.0%) | 195 (5.8%) | 990 (42.4%) | 845 (45.2%) | 1,260 (16.0%) | 1,040 (19.8%) |
| **Region** | | | | | | | | | | |
| East of England | 51,955 (21.7%) | 35,665 (22.7%) | 2,210 (21.4%) | 1,545 (22.1%) | 1,365 (24.6%) | 785 (23.2%) | 420 (18.0%) | 530 (28.3%) | 1,790 (22.7%) | 1,315 (25.0%) |
| Midlands | 57,865 (24.2%) | 37,655 (24.0%) | 2,440 (23.6%) | 1,590 (22.8%) | 1,240 (22.4%) | 725 (21.4%) | 645 (27.6%) | 370 (19.8%) | 1,885 (23.9%) | 1,090 (20.7%) |
| London | 4,805 (2.0%) | 4,925 (3.1%) | 360 (3.5%) | 360 (5.2%) | 255 (4.6%) | 320 (9.5%) | 170 (7.3%) | 115 (6.1%) | 425 (5.4%) | 435 (8.3%) |
| North East and Yorkshire | 45,570 (19.0%) | 28,265 (18.0%) | 2,045 (19.8%) | 1,305 (18.7%) | 865 (15.6%) | 525 (15.5%) | 435 (18.6%) | 330 (17.6%) | 1,300 (16.5%) | 855 (16.3%) |
| North West | 22,825 (9.5%) | 13,360 (8.5%) | 1,040 (10.1%) | 645 (9.2%) | 565 (10.2%) | 335 (9.9%) | 165 (7.1%) | 195 (10.4%) | 725 (9.2%) | 530 (10.1%) |
| South East | 12,635 (5.3%) | 8,380 (5.3%) | 565 (5.5%) | 375 (5.4%) | 380 (6.9%) | 165 (4.9%) | 160 (6.9%) | 100 (5.3%) | 540 (6.9%) | 265 (5.0%) |
| South West | 43,725 (18.3%) | 28,710 (18.3%) | 1,660 (16.1%) | 1,165 (16.7%) | 870 (15.7%) | 525 (15.5%) | 345 (14.8%) | 240 (12.8%) | 1,210 (15.4%) | 765 (14.6%) |
| **JCVI priority group** | | | | | | | | | | |
| 3 (75+) | 77,190 (32.2%) | 69,290 (44.1%) | 3,380 (32.7%) | 2,965 (42.4%) | 225 (4.1%) | 220 (6.5%) | 355 (15.2%) | 380 (20.3%) | 575 (7.3%) | 600 (11.4%) |
| 4 (70+ or clinically extremely vulnerable) | 91,610 (38.3%) | 50,965 (32.5%) | 4,750 (46.0%) | 2,645 (37.9%) | 5,265 (95.0%) | 3,125 (92.3%) | 1,945 (83.3%) | 1,440 (77.0%) | 7,210 (91.6%) | 4,565 (86.9%) |
| 5 (65+) | 33,135 (13.8%) | 17,110 (10.9%) | 885 (8.6%) | 505 (7.2%) | ≤10 | 0 (0.0%) | ≤10 | 15 (0.8%) | 15 (0.2%) | 15 (0.3%) |
| 6 (16–65 and clinically vulnerable) | 37,440 (15.6%) | 19,595 (12.5%) | 1,305 (12.6%) | 870 (12.5%) | 45 (0.8%) | 35 (1.0%) | 30 (1.3%) | 40 (2.1%) | 75 (1.0%) | 75 (1.4%) |

Data are n (%) after rounding to the nearest 5. AZ, AZD1222 (AstraZeneca); BNT, BNT162b2 (Pfizer/BioNTech). CKD, chronic kidney disease; JCVI, Joint Committee on Vaccination and Immunisation; [N], absent in all individuals by definition; RRT, renal replacement therapy.

**Table S7. Baseline characteristics of three-dose cohort subgroups.**

| Characteristic | CKD3 | | CKD4–5 | | RRT (transplant) | | RRT (dialysis) | | RRT (combined) | |
|---|---|---|---|---|---|---|---|---|---|---|
| | **AZ–AZ N = 205,500** | **BNT–BNT N = 145,815** | **AZ–AZ N = 9,605** | **BNT–BNT N = 6,885** | **AZ–AZ N = 4,445** | **BNT–BNT N = 2,885** | **AZ–AZ N = 1,785** | **BNT–BNT N = 1,480** | **AZ–AZ N = 6,230** | **BNT–BNT N = 4,365** |
| **Age** | | | | | | | | | | |
| 16–64 | 32,055 (15.7%) | 19,235 (13.2%) | 2,020 (21.0%) | 1,340 (19.5%) | 3,245 (73.0%) | 2,040 (70.7%) | 930 (52.1%) | 730 (49.3%) | 4,180 (67.1%) | 2,770 (63.5%) |
| 65–69 | 31,935 (15.6%) | 18,030 (12.4%) | 1,320 (13.7%) | 770 (11.2%) | 575 (12.9%) | 335 (11.6%) | 270 (15.1%) | 200 (13.5%) | 845 (13.6%) | 535 (12.3%) |
| 70–74 | 69,160 (33.8%) | 41,000 (28.1%) | 2,860 (29.8%) | 1,630 (23.7%) | 430 (9.7%) | 320 (11.1%) | 280 (15.7%) | 250 (16.9%) | 715 (11.5%) | 570 (13.1%) |
| 75–79 | 71,350 (34.9%) | 67,550 (46.3%) | 3,405 (35.5%) | 3,150 (45.8%) | 190 (4.3%) | 190 (6.6%) | 305 (17.1%) | 295 (19.9%) | 495 (7.9%) | 490 (11.2%) |
| **Sex** | | | | | | | | | | |
| Female | 109,110 (53.4%) | 77,940 (53.5%) | 4,135 (43.1%) | 2,980 (43.3%) | 1,685 (37.9%) | 1,085 (37.6%) | 660 (37.0%) | 515 (34.8%) | 2,345 (37.6%) | 1,605 (36.8%) |
| Male | 95,390 (46.6%) | 67,875 (46.5%) | 5,465 (56.9%) | 3,905 (56.7%) | 2,760 (62.1%) | 1,800 (62.4%) | 1,125 (63.0%) | 960 (64.9%) | 3,885 (62.4%) | 2,760 (63.2%) |
| **Ethnicity** | | | | | | | | | | |
| White | 193,560 (94.7%) | 138,245 (94.8%) | 8,700 (90.6%) | 6,225 (90.4%) | 3,830 (86.2%) | 2,430 (84.2%) | 1,410 (79.0%) | 1,225 (82.8%) | 5,240 (84.1%) | 3,650 (83.6%) |
| Black | 2,835 (1.4%) | 1,830 (1.3%) | 180 (1.9%) | 130 (1.9%) | 105 (2.4%) | 80 (2.8%) | 100 (5.6%) | 60 (4.1%) | 205 (3.3%) | 140 (3.2%) |
| South Asian | 5,995 (2.9%) | 4,260 (2.9%) | 570 (5.9%) | 435 (6.3%) | 400 (9.0%) | 290 (10.1%) | 225 (12.6%) | 155 (10.5%) | 625 (10.0%) | 450 (10.3%) |
| Mixed | 865 (0.4%) | 650 (0.4%) | 60 (0.6%) | 40 (0.6%) | 35 (0.8%) | 20 (0.7%) | 15 (0.8%) | 15 (1.0%) | 50 (0.8%) | 40 (0.9%) |
| Other | 1,245 (0.6%) | 830 (0.6%) | 95 (1.0%) | 50 (0.7%) | 70 (1.6%) | 65 (2.3%) | 40 (2.2%) | 20 (1.4%) | 110 (1.8%) | 85 (1.9%) |
| **Index of multiple deprivation quintile** | | | | | | | | | | |
| 1 most deprived | 32,555 (15.9%) | 22,795 (15.6%) | 1,905 (19.8%) | 1,310 (19.0%) | 790 (17.8%) | 485 (16.8%) | 505 (28.3%) | 340 (23.0%) | 1,295 (20.8%) | 825 (18.9%) |
| 2 | 38,550 (18.9%) | 27,450 (18.8%) | 1,970 (20.5%) | 1,435 (20.8%) | 875 (19.7%) | 585 (20.3%) | 430 (24.1%) | 340 (23.0%) | 1,305 (20.9%) | 925 (21.2%) |
| 3 | 46,885 (22.9%) | 33,490 (23.0%) | 2,175 (22.6%) | 1,595 (23.2%) | 985 (22.2%) | 650 (22.5%) | 335 (18.8%) | 310 (20.9%) | 1,320 (21.2%) | 965 (22.1%) |
| 4 | 45,150 (22.1%) | 32,405 (22.2%) | 1,900 (19.8%) | 1,360 (19.8%) | 950 (21.4%) | 595 (20.6%) | 270 (15.1%) | 270 (18.2%) | 1,220 (19.6%) | 865 (19.8%) |
| 5 least deprived | 41,360 (20.2%) | 29,670 (20.3%) | 1,655 (17.2%) | 1,180 (17.1%) | 845 (19.0%) | 570 (19.8%) | 245 (13.7%) | 215 (14.5%) | 1,090 (17.5%) | 785 (18.0%) |
| **Setting** | | | | | | | | | | |
| Urban city or town | 109,690 (53.6%) | 77,000 (52.8%) | 4,965 (51.7%) | 3,520 (51.1%) | 2,470 (55.6%) | 1,560 (54.1%) | 850 (47.6%) | 835 (56.4%) | 3,320 (53.3%) | 2,395 (54.9%) |
| Urban conurbation | 40,295 (19.7%) | 29,005 (19.9%) | 2,315 (24.1%) | 1,685 (24.5%) | 960 (21.6%) | 650 (22.5%) | 600 (33.6%) | 335 (22.6%) | 1,560 (25.0%) | 985 (22.6%) |
| Rural | 54,515 (26.7%) | 39,815 (27.3%) | 2,320 (24.2%) | 1,675 (24.3%) | 1,015 (22.8%) | 675 (23.4%) | 335 (18.8%) | 310 (20.9%) | 1,350 (21.7%) | 985 (22.6%) |
| **Kidney disease** | | | | | | | | | | |
| CKD3a | 164,575 (80.5%) | 115,730 (79.4%) | [N] | [N] | [N] | [N] | [N] | [N] | [N] | [N] |
| CKD3b | 39,920 (19.5%) | 30,085 (20.6%) | [N] | [N] | [N] | [N] | [N] | [N] | [N] | [N] |
| CKD4–5 | [N] | [N] | 9,605 (100.0%) | 6,885 (100.0%) | [N] | [N] | [N] | [N] | [N] | [N] |
| RRT (dialysis) | [N] | [N] | [N] | [N] | [N] | [N] | 1,785 (100.0%) | 1,480 (100.0%) | 1,785 (28.7%) | 1,480 (33.9%) |
| RRT (transplant) | [N] | [N] | [N] | [N] | 4,445 (100.0%) | 2,885 (100.0%) | [N] | [N] | 4,445 (71.3%) | 2,885 (66.1%) |
| **Primary care coding of kidney disease** | | | | | | | | | | |
| CKD3–5 | 108,210 (52.9%) | 80,545 (55.2%) | 8,295 (86.4%) | 6,115 (88.8%) | 3,405 (76.6%) | 2,250 (78.0%) | 1,530 (85.7%) | 1,255 (84.8%) | 4,935 (79.2%) | 3,505 (80.3%) |
| Dialysis code | [N] | [N] | [N] | [N] | 2,890 (65.0%) | 1,840 (63.8%) | 1,495 (83.8%) | 1,230 (83.1%) | 4,385 (70.4%) | 3,070 (70.3%) |
| Kidney transplant code | [N] | [N] | [N] | [N] | 4,330 (97.4%) | 2,800 (97.1%) | 385 (21.6%) | 300 (20.3%) | 4,715 (75.7%) | 3,100 (71.0%) |
| **Morbidities** | | | | | | | | | | |
| Immunosuppression | 11,885 (5.8%) | 8,420 (5.8%) | 790 (8.2%) | 515 (7.5%) | 1,710 (38.5%) | 1,125 (39.0%) | 225 (12.6%) | 185 (12.5%) | 1,935 (31.1%) | 1,310 (30.0%) |
| Severe obesity | 12,130 (5.9%) | 8,550 (5.9%) | 820 (8.5%) | 620 (9.0%) | 110 (2.5%) | 75 (2.6%) | 125 (7.0%) | 110 (7.4%) | 235 (3.8%) | 185 (4.2%) |
| Diabetes | 55,570 (27.2%) | 40,710 (27.9%) | 4,740 (49.3%) | 3,470 (50.4%) | 1,405 (31.6%) | 900 (31.2%) | 740 (41.5%) | 590 (39.9%) | 2,145 (34.4%) | 1,495 (34.2%) |
| Chronic respiratory disease (inc. asthma) | 23,155 (11.3%) | 16,675 (11.4%) | 1,340 (14.0%) | 920 (13.4%) | 335 (7.5%) | 225 (7.8%) | 190 (10.6%) | 175 (11.8%) | 525 (8.4%) | 400 (9.2%) |
| Chronic heart disease | 73,730 (36.1%) | 54,985 (37.7%) | 4,540 (47.3%) | 3,315 (48.1%) | 1,370 (30.8%) | 935 (32.4%) | 880 (49.3%) | 765 (51.7%) | 2,250 (36.1%) | 1,700 (38.9%) |
| Chronic liver disease | 9,240 (4.5%) | 6,425 (4.4%) | 565 (5.9%) | 400 (5.8%) | 225 (5.1%) | 155 (5.4%) | 105 (5.9%) | 80 (5.4%) | 325 (5.2%) | 235 (5.4%) |
| Asplenia | 1,965 (1.0%) | 1,395 (1.0%) | 130 (1.4%) | 60 (0.9%) | 70 (1.6%) | 40 (1.4%) | 30 (1.7%) | 15 (1.0%) | 105 (1.7%) | 50 (1.1%) |
| Haematologic cancer | 4,080 (2.0%) | 3,075 (2.1%) | 320 (3.3%) | 215 (3.1%) | 80 (1.8%) | 75 (2.6%) | 55 (3.1%) | 50 (3.4%) | 135 (2.2%) | 125 (2.9%) |
| Organ transplant (non-kidney) | 775 (0.4%) | 395 (0.3%) | 95 (1.0%) | 55 (0.8%) | 0 (0.0%) | 0 (0.0%) | 15 (0.8%) | ≤10 | 15 (0.2%) | ≤10 |
| Chronic neurological disease | 21,900 (10.7%) | 15,965 (10.9%) | 1,320 (13.7%) | 955 (13.9%) | 420 (9.4%) | 280 (9.7%) | 240 (13.4%) | 215 (14.5%) | 660 (10.6%) | 495 (11.3%) |
| Learning disability | 765 (0.4%) | 390 (0.3%) | 55 (0.6%) | 35 (0.5%) | 50 (1.1%) | 50 (1.7%) | 15 (0.8%) | 15 (1.0%) | 65 (1.0%) | 60 (1.4%) |

| | | | | | | | | | | |
|---|---|---|---|---|---|---|---|---|---|---|
| Severe mental illness | 2,730 (1.3%) | 1,695 (1.2%) | 205 (2.1%) | 115 (1.7%) | 40 (0.9%) | 25 (0.9%) | 45 (2.5%) | 20 (1.4%) | 85 (1.4%) | 45 (1.0%) |
| Clinically extremely vulnerable | 39,220 (19.2%) | 28,530 (19.6%) | 4,210 (43.8%) | 2,940 (42.7%) | 4,390 (98.8%) | 2,855 (99.0%) | 1,755 (98.3%) | 1,425 (96.3%) | 6,145 (98.6%) | 4,285 (98.2%) |
| **Prior SARS-CoV-2 infection** | 7,120 (3.5%) | 3,850 (2.6%) | 495 (5.2%) | 260 (3.8%) | 285 (6.4%) | 170 (5.9%) | 310 (17.4%) | 190 (12.8%) | 595 (9.6%) | 360 (8.2%) |
| **No. of SARS-CoV-2 tests in 90-day pre-vaccination window** | | | | | | | | | | |
| 0 | 160,355 (78.4%) | 114,465 (78.5%) | 6,970 (72.6%) | 4,995 (72.5%) | 3,005 (67.6%) | 1,950 (67.6%) | 425 (23.8%) | 365 (24.7%) | 3,430 (55.1%) | 2,315 (53.0%) |
| 1 | 20,700 (10.1%) | 14,675 (10.1%) | 1,080 (11.2%) | 785 (11.4%) | 610 (13.7%) | 390 (13.5%) | 130 (7.3%) | 140 (9.5%) | 740 (11.9%) | 525 (12.0%) |
| 2 | 7,810 (3.8%) | 5,435 (3.7%) | 470 (4.9%) | 325 (4.7%) | 270 (6.1%) | 165 (5.7%) | 95 (5.3%) | 75 (5.1%) | 365 (5.9%) | 240 (5.5%) |
| 3+ | 15,635 (7.6%) | 11,245 (7.7%) | 1,085 (11.3%) | 780 (11.3%) | 555 (12.5%) | 385 (13.3%) | 1,140 (63.9%) | 900 (60.8%) | 1,700 (27.3%) | 1,285 (29.4%) |
| **Region** | | | | | | | | | | |
| East of England | 43,110 (21.1%) | 33,305 (22.8%) | 1,945 (20.2%) | 1,490 (21.6%) | 1,115 (25.1%) | 685 (23.7%) | 335 (18.8%) | 400 (27.0%) | 1,455 (23.4%) | 1,085 (24.9%) |
| Midlands | 48,285 (23.6%) | 34,105 (23.4%) | 2,285 (23.8%) | 1,595 (23.2%) | 1,015 (22.8%) | 635 (22.0%) | 510 (28.6%) | 305 (20.6%) | 1,525 (24.5%) | 940 (21.5%) |
| London | 3,990 (2.0%) | 4,790 (3.3%) | 315 (3.3%) | 365 (5.3%) | 175 (3.9%) | 265 (9.2%) | 115 (6.4%) | 95 (6.4%) | 290 (4.7%) | 360 (8.2%) |
| North East and Yorkshire | 39,975 (19.5%) | 26,625 (18.3%) | 1,955 (20.4%) | 1,255 (18.2%) | 615 (13.8%) | 410 (14.2%) | 305 (17.1%) | 260 (17.6%) | 920 (14.8%) | 665 (15.2%) |
| North West | 19,615 (9.6%) | 12,555 (8.6%) | 935 (9.7%) | 620 (9.0%) | 445 (10.0%) | 285 (9.9%) | 130 (7.3%) | 150 (10.1%) | 575 (9.2%) | 435 (10.0%) |
| South East | 12,265 (6.0%) | 8,355 (5.7%) | 585 (6.1%) | 410 (6.0%) | 320 (7.2%) | 155 (5.4%) | 115 (6.4%) | 85 (5.7%) | 435 (7.0%) | 240 (5.5%) |
| South West | 37,260 (18.2%) | 26,075 (17.9%) | 1,580 (16.4%) | 1,150 (16.7%) | 750 (16.9%) | 455 (15.8%) | 280 (15.7%) | 185 (12.5%) | 1,030 (16.5%) | 640 (14.7%) |
| **JCVI priority group** | | | | | | | | | | |
| 3 (75+) | 71,350 (34.9%) | 67,550 (46.3%) | 3,405 (35.5%) | 3,150 (45.8%) | 190 (4.3%) | 190 (6.6%) | 305 (17.1%) | 295 (19.9%) | 495 (7.9%) | 490 (11.2%) |
| 4 (70+ or clinically extremely vulnerable) | 82,815 (40.5%) | 48,880 (33.5%) | 4,510 (47.0%) | 2,625 (38.1%) | 4,205 (94.6%) | 2,670 (92.5%) | 1,465 (82.1%) | 1,145 (77.4%) | 5,670 (91.0%) | 3,815 (87.4%) |
| 5 (65+) | 25,815 (12.6%) | 14,695 (10.1%) | 725 (7.5%) | 430 (6.2%) | ≤10 | 0 (0.0%) | ≤10 | ≤10 | ≤10 | ≤10 |
| 6 (16–65 and clinically vulnerable) | 24,520 (12.0%) | 14,690 (10.1%) | 965 (10.0%) | 680 (9.9%) | 45 (1.0%) | 25 (0.9%) | 15 (0.8%) | 25 (1.7%) | 60 (1.0%) | 50 (1.1%) |

Data are n (%) after rounding to the nearest 5. Non-zero rounded counts of ≤10 redacted. AZ, AZD1222 (AstraZeneca); BNT, BNT162b2 (Pfizer/BioNTech). CKD, chronic kidney disease; JCVI, Joint Committee on Vaccination and Immunisation; [N], absent in all individuals by definition; RRT, renal replacement therapy.

**Table S8. Incidence rates and hazard ratios for two-dose cohort.**

| | BNT–BNT | | | | | | AZ–AZ | | | | | | | Hazard ratio (95% CI) | | | |
|---|---|---|---|---|---|---|---|---|---|---|---|---|---|---|---|---|---|
| Period (days) | N | N events | Person-years | Rate (per 1,000 p-y) | N, matched | N events, matched | N | N events | Person-years | Rate (per 1,000 p-y) | N, matched | N events, matched | Incidence rate ratio (95% CI) | Unadjusted | Region/date adjusted | Fully adjusted | Matched |
| **Positive SARS-CoV-2 test** | | | | | | | | | | | | | | | | | |
| 1-14 | 169,205 | 25 | 6,479 | 4.2 | 130,765 | 15 | 257,580 | 40 | 9,867 | 4.3 | 130,765 | ≤10 | 1.02 (0.62-1.81) | 1.02 (0.63-1.66) | 0.86 (0.53-1.4) | 0.92 (0.57-1.5) | [R] |
| 15-70 | 168,930 | 155 | 25,831 | 6.0 | 130,565 | 100 | 257,255 | 590 | 39,334 | 15.0 | 130,615 | 165 | 2.52 (2.09-3.00) | 2.52 (2.11-3.01) | 2.14 (1.8-2.56) | 2.28 (1.91-2.72) | 1.63 (1.28-2.09) |
| 71-126 | 167,780 | 1,025 | 25,535 | 40.1 | 130,035 | 760 | 255,530 | 2,665 | 38,879 | 68.6 | 129,910 | 1,150 | 1.71 (1.59-1.84) | 1.71 (1.59-1.84) | 1.44 (1.34-1.55) | 1.51 (1.4-1.62) | 1.52 (1.39-1.66) |
| 127-182 | 165,020 | 1,865 | 24,964 | 74.6 | 128,705 | 1,440 | 250,845 | 4,035 | 36,933 | 109.2 | 128,130 | 1,950 | 1.46 (1.38-1.55) | 1.47 (1.39-1.55) | 1.27 (1.21-1.35) | 1.33 (1.26-1.41) | 1.36 (1.27-1.46) |
| **1-182** | **169,205** | **3,070** | **82,809** | **37.1** | **130,765** | **2,315** | **257,580** | **7,335** | **125,013** | **58.7** | **130,765** | **3,270** | **1.58 (1.52-1.65)** | **1.6 (1.53-1.67)** | **1.37 (1.31-1.43)** | **1.43 (1.37-1.5)** | **1.42 (1.35-1.5)** |
| **COVID-19-related hospitalisation** | | | | | | | | | | | | | | | | | |
| 1-14 | 169,205 | ≤10 | 6,479 | [R] | 130,765 | ≤10 | 257,580 | ≤10 | 9,867 | [R] | 130,765 | 0 | [R] | [R] | [R] | [R] | [R] |
| 15-70 | 168,950 | 25 | 25,840 | 1.0 | 130,570 | 15 | 257,290 | 65 | 39,365 | 1.6 | 130,620 | 25 | 1.54 (1.06-2.83) | 1.53 (0.98-2.4) | 1.49 (0.95-2.34) | 1.47 (0.94-2.31) | 1.92 (0.98-3.76) |
| 71-126 | 167,925 | 165 | 25,606 | 6.5 | 130,135 | 115 | 256,095 | 470 | 39,108 | 12.0 | 130,055 | 205 | 1.84 (1.56-2.24) | 1.83 (1.54-2.19) | 1.79 (1.5-2.14) | 1.76 (1.47-2.1) | 1.8 (1.43-2.26) |
| 127-182 | 166,000 | 280 | 25,220 | 11.0 | 129,445 | 210 | 253,570 | 650 | 37,542 | 17.3 | 129,210 | 305 | 1.57 (1.35-1.80) | 1.58 (1.37-1.81) | 1.55 (1.34-1.78) | 1.52 (1.31-1.75) | 1.45 (1.22-1.73) |
| **1-182** | **169,205** | **475** | **83,146** | **5.7** | **130,765** | **345** | **257,580** | **1,185** | **125,883** | **9.4** | **130,765** | **540** | **1.65 (1.48-1.84)** | **1.65 (1.49-1.84)** | **1.62 (1.45-1.81)** | **1.59 (1.43-1.77)** | **1.57 (1.38-1.8)** |
| **COVID-19-related death** | | | | | | | | | | | | | | | | | |
| 1-14 | 169,205 | 0 | 6,479 | -- | 130,765 | 0 | 257,580 | 0 | 9,868 | -- | 130,765 | 0 | [R] | [R] | [R] | [R] | [R] |
| 15-70 | 168,955 | ≤10 | 25,842 | [R] | 130,575 | 0 | 257,295 | ≤10 | 39,369 | [R] | 130,625 | 0 | [R] | [R] | [R] | [R] | [R] |
| 71-126 | 167,955 | 30 | 25,619 | 1.1 | 130,150 | 15 | 256,155 | 75 | 39,142 | 1.9 | 130,085 | 30 | 1.67 (1.06-2.59) | 1.67 (1.09-2.57) | 1.63 (1.06-2.51) | 1.59 (1.03-2.44) | 1.88 (1.05-3.39) |
| 127-182 | 166,165 | 60 | 25,260 | 2.5 | 129,555 | 45 | 254,025 | 135 | 37,644 | 3.6 | 129,405 | 65 | 1.45 (1.11-2.08) | 1.45 (1.07-1.96) | 1.44 (1.06-1.96) | 1.41 (1.03-1.92) | 1.42 (0.97-2.09) |
| **1-182** | **169,205** | **95** | **83,201** | **1.1** | **130,765** | **65** | **257,580** | **210** | **126,023** | **1.7** | **130,765** | **95** | **1.46 (1.14-1.88)** | **1.49 (1.17-1.91)** | **1.48 (1.15-1.89)** | **1.44 (1.12-1.85)** | **1.54 (1.12-2.12)** |
| **Non-COVID-19 death** | | | | | | | | | | | | | | | | | |
| 1-14 | 169,205 | 55 | 6,479 | 8.2 | 130,765 | 40 | 257,580 | 80 | 9,868 | 8.0 | 130,765 | 35 | 0.98 (0.67-1.37) | 0.98 (0.69-1.39) | 1.05 (0.74-1.48) | 1 (0.7-1.41) | 0.86 (0.55-1.34) |
| 15-70 | 168,955 | 390 | 25,842 | 15.1 | 130,575 | 270 | 257,295 | 655 | 39,369 | 16.6 | 130,625 | 330 | 1.10 (0.97-1.25) | 1.1 (0.97-1.25) | 1.17 (1.03-1.33) | 1.12 (0.99-1.27) | 1.23 (1.05-1.45) |
| 71-126 | 167,955 | 525 | 25,619 | 20.5 | 130,150 | 385 | 256,155 | 890 | 39,142 | 22.7 | 130,085 | 435 | 1.11 (1.00-1.24) | 1.11 (0.99-1.23) | 1.18 (1.06-1.32) | 1.13 (1.01-1.26) | 1.14 (0.99-1.3) |
| 127-182 | 166,165 | 575 | 25,260 | 22.8 | 129,555 | 400 | 254,025 | 885 | 37,644 | 23.5 | 129,405 | 430 | 1.03 (0.93-1.15) | 1.03 (0.93-1.15) | 1.08 (0.97-1.2) | 1.04 (0.93-1.16) | 1.07 (0.94-1.23) |
| **1-182** | **169,205** | **1,545** | **83,201** | **18.6** | **130,765** | **1,095** | **257,580** | **2,505** | **126,023** | **19.9** | **130,765** | **1,230** | **1.07 (1.00-1.14)** | **1.07 (1.01-1.14)** | **1.14 (1.06-1.21)** | **1.09 (1.02-1.16)** | **1.13 (1.04-1.22)** |

Counts are rounded to the nearest 5, and non-zero rounded counts of ≤10 redacted. AZ, AZD1222 (AstraZeneca); BNT, BNT162b2 (Pfizer/BioNTech); p-y, person-year; [R], redacted due to low event counts in one or both vaccine groups.

**Table S9. Incidence rates and hazard ratios for kidney disease subgroups (two-dose cohort).**

| | BNT–BNT | | | | AZ–AZ | | | | | |
|---|---|---|---|---|---|---|---|---|---|---|
| **Subgroup** | **N** | **N events** | **Person-years** | **Rate (per 1,000 p-y)** | **N** | **N events** | **Person-years** | **Rate (per 1,000 p-y)** | **Incidence rate ratio (95% CI)** | **Fully adjusted hazard ratio (95% CI)** |
| **Positive SARS-CoV-2 test** | | | | | | | | | | |
| **All** | **169,205** | **3,070** | **82,809** | **37.1** | **257,580** | **7,335** | **125,013** | **58.7** | **1.58 (1.52-1.65)** | **1.43 (1.37-1.50)** |
| CKD3 | 156,965 | 2,665 | 76,938 | 34.6 | 239,380 | 6,505 | 116,352 | 55.9 | 1.61 (1.54-1.69) | 1.45 (1.39-1.52) |
| CKD4–5 | 6,985 | 165 | 3,380 | 48.8 | 10,325 | 315 | 4,960 | 63.5 | 1.30 (1.07-1.58) | 1.27 (1.04-1.54) |
| RRT (transplant) | 3,385 | 150 | 1,602 | 93.6 | 5,540 | 375 | 2,609 | 143.7 | 1.54 (1.27-1.87) | 1.41 (1.16-1.71) |
| RRT (dialysis) | 1,870 | 90 | 889 | 101.2 | 2,335 | 140 | 1,092 | 128.2 | 1.27 (0.97-1.67) | 1.16 (0.87-1.54) |
| RRT (combined) | 5,255 | 240 | 2,491 | 96.3 | 7,875 | 510 | 3,701 | 137.8 | 1.43 (1.22-1.67) | 1.35 (1.15-1.58) |
| **COVID-19-related hospitalisation** | | | | | | | | | | |
| **All** | **169,205** | **475** | **83,146** | **5.7** | **257,580** | **1,185** | **125,883** | **9.4** | **1.65 (1.48-1.84)** | **1.59 (1.43-1.77)** |
| CKD3 | 156,965 | 350 | 77,237 | 4.5 | 239,380 | 885 | 117,142 | 7.6 | 1.67 (1.47-1.89) | 1.59 (1.40-1.81) |
| CKD4–5 | 6,985 | 45 | 3,396 | 13.3 | 10,325 | 110 | 4,989 | 22.1 | 1.66 (1.17-2.41) | 1.66 (1.16-2.38) |
| RRT (transplant) | 3,385 | 60 | 1,616 | 37.1 | 5,540 | 145 | 2,646 | 54.8 | 1.48 (1.09-2.03) | 1.39 (1.01-1.90) |
| RRT (dialysis) | 1,870 | 20 | 897 | 22.3 | 2,335 | 45 | 1,106 | 40.7 | 1.83 (1.06-3.26) | 1.73 (1.00-3.00) |
| RRT (combined) | 5,255 | 80 | 2,513 | 31.8 | 7,875 | 190 | 3,752 | 50.6 | 1.59 (1.22-2.09) | 1.47 (1.12-1.92) |
| **COVID-19-related death** | | | | | | | | | | |
| **All** | **169,205** | **95** | **83,201** | **1.1** | **257,580** | **210** | **126,023** | **1.7** | **1.46 (1.14-1.88)** | **1.44 (1.12-1.85)** |
| CKD3 | 156,965 | 65 | 77,277 | 0.8 | 239,380 | 145 | 117,246 | 1.2 | 1.48 (1.09-2.00) | 1.46 (1.08-1.98) |
| CKD4–5 | 6,985 | ≤10 | 3,401 | [R] | 10,325 | 25 | 5,002 | 5.0 | [R] | [R] |
| RRT (transplant) | 3,385 | 20 | 1,623 | 12.3 | 5,540 | 30 | 2,664 | 11.3 | 0.91 (0.50-1.70) | 1.16 (0.63-2.13) |
| RRT (dialysis) | 1,870 | ≤10 | 900 | [R] | 2,335 | ≤10 | 1,111 | [R] | [R] | [R] |
| RRT (combined) | 5,255 | 25 | 2,523 | 9.9 | 7,875 | 45 | 3,775 | 11.9 | 1.20 (0.72-2.05) | 1.34 (0.79-2.28) |
| **Non-COVID-19 death** | | | | | | | | | | |
| **All** | **169,205** | **1,545** | **83,201** | **18.6** | **257,580** | **2,505** | **126,023** | **19.9** | **1.07 (1.00-1.14)** | **1.09 (1.02-1.16)** |
| CKD3 | 156,965 | 1,245 | 77,277 | 16.1 | 239,380 | 2,075 | 117,246 | 17.7 | 1.10 (1.02-1.18) | 1.11 (1.03-1.19) |
| CKD4–5 | 6,985 | 185 | 3,401 | 54.4 | 10,325 | 275 | 5,002 | 55.0 | 1.01 (0.84-1.22) | 0.99 (0.82-1.20) |
| RRT (transplant) | 3,385 | 30 | 1,623 | 18.5 | 5,540 | 50 | 2,664 | 18.8 | 1.02 (0.63-1.65) | 1.05 (0.65-1.70) |
| RRT (dialysis) | 1,870 | 85 | 900 | 94.4 | 2,335 | 110 | 1,111 | 99.0 | 1.05 (0.78-1.41) | 1.17 (0.87-1.59) |
| RRT (combined) | 5,255 | 115 | 2,523 | 45.6 | 7,875 | 160 | 3,775 | 42.4 | 0.93 (0.73-1.19) | 1.10 (0.85-1.41) |

Counts are rounded to the nearest 5, and non-zero rounded counts of ≤10 redacted. AZ, AZD1222 (AstraZeneca); BNT, BNT162b2 (Pfizer/BioNTech); CKD, chronic kidney disease; p-y, person-year; [R], redacted due to low event counts in one or both vaccine groups; RRT, renal replacement therapy.

**Table S10. Incidence rates and hazard ratios for three-dose cohort.**

| | BNT–BNT–BNT | | | | | | AZ–AZ–BNT | | | | | | | Hazard ratio (95% CI) | | | |
|---|---|---|---|---|---|---|---|---|---|---|---|---|---|---|---|---|---|
| Period (days) | N | N events | Person-years | Rate (per 1,000 p-y) | N, matched | N events, matched | N | N events | Person-years | Rate (per 1,000 p-y) | N, matched | N events, matched | Incidence rate ratio (95% CI) | Unadjusted | Region/date adjusted | Fully adjusted | Matched |
| **Positive SARS-CoV-2 test** | | | | | | | | | | | | | | | | | |
| 1-14 | 157,065 | 535 | 6,008 | 88.9 | 127,345 | 390 | 220,330 | 1,000 | 8,422 | 119.0 | 127,345 | 495 | 1.34 (1.20-1.48) | 1.34 (1.21-1.49) | 1.28 (1.15-1.42) | 1.25 (1.13-1.39) | 1.26 (1.11-1.44) |
| 15-70 | 156,370 | 3,065 | 23,766 | 129.0 | 126,835 | 2320 | 219,080 | 4,850 | 33,256 | 145.9 | 126,720 | 2340 | 1.13 (1.08-1.18) | 1.13 (1.08-1.18) | 1.08 (1.03-1.13) | 1.06 (1.01-1.11) | 1.01 (0.95-1.07) |
| 71-126 | 152,635 | 4,960 | 22,544 | 219.9 | 124,050 | 3590 | 213,340 | 6,180 | 31,505 | 196.2 | 123,940 | 3480 | 0.89 (0.86-0.93) | 0.91 (0.88-0.95) | 0.88 (0.84-0.91) | 0.86 (0.83-0.90) | 0.97 (0.93-1.02) |
| 127-182 | 139,880 | 3,305 | 18,973 | 174.1 | 115,880 | 2525 | 195,155 | 3,425 | 25,803 | 132.7 | 115,650 | 2325 | 0.76 (0.73-0.80) | 0.97 (0.92-1.02) | 0.96 (0.92-1.01) | 0.95 (0.90-0.99) | 0.94 (0.89-0.99) |
| **1-182** | **157,065** | **11,860** | **71,290** | **166.4** | **127,345** | **8,820** | **220,330** | **15,460** | **98,986** | **156.2** | **127,345** | **8,640** | **0.94 (0.92-0.96)** | **1.01 (0.98-1.03)** | **0.97 (0.95-1.00)** | **0.96 (0.93-0.98)** | **0.99 (0.96-1.02)** |
| **COVID-19-related hospitalisation** | | | | | | | | | | | | | | | | | |
| 1-14 | 157,065 | 50 | 6,016 | 8.6 | 127,345 | 35 | 220,330 | 105 | 8,438 | 12.7 | 127,345 | 50 | 1.47 (1.06-2.14) | 1.47 (1.05-2.04) | 1.46 (1.05-2.03) | 1.45 (1.04-2.03) | 1.53 (0.99-2.36) |
| 15-70 | 156,850 | 265 | 23,985 | 11.0 | 127,190 | 165 | 219,970 | 435 | 33,634 | 13.0 | 127,160 | 185 | 1.18 (1.00-1.37) | 1.18 (1.01-1.38) | 1.19 (1.02-1.38) | 1.18 (1.01-1.38) | 1.13 (0.92-1.40) |
| 71-126 | 155,920 | 515 | 23,400 | 22.1 | 126,560 | 340 | 218,640 | 775 | 32,735 | 23.6 | 126,535 | 380 | 1.07 (0.96-1.20) | 1.07 (0.96-1.19) | 1.07 (0.96-1.20) | 1.07 (0.96-1.20) | 1.11 (0.96-1.29) |
| 127-182 | 147,390 | 570 | 20,360 | 27.9 | 121,525 | 430 | 205,630 | 645 | 27,621 | 23.3 | 121,245 | 345 | 0.83 (0.74-0.94) | 0.83 (0.74-0.93) | 0.84 (0.75-0.94) | 0.84 (0.75-0.94) | 0.80 (0.70-0.92) |
| **1-182** | **157,065** | **1400** | **73,762** | **19.0** | **127,345** | **970** | **220,330** | **1,960** | **102,428** | **19.1** | **127,345** | **960** | **1.01 (0.94-1.08)** | **1.01 (0.94-1.08)** | **1.01 (0.95-1.09)** | **1.01 (0.95-1.09)** | **0.99 (0.91-1.08)** |
| **COVID-19-related death** | | | | | | | | | | | | | | | | | |
| 1-14 | 157,065 | ≤10 | 6,017 | [R] | 127,345 | 0 | 220,330 | ≤10 | 8,440 | [R] | 127,345 | ≤10 | [R] | [R] | [R] | [R] | [R] |
| 15-70 | 156,900 | 35 | 24,007 | 1.5 | 127,225 | 20 | 220,070 | 50 | 33,674 | 1.5 | 127,210 | 15 | 1.05 (0.65-1.62) | 1.06 (0.69-1.63) | 1.19 (0.77-1.83) | 1.15 (0.75-1.77) | 0.79 (0.40-1.55) |
| 71-126 | 156,190 | 50 | 23,473 | 2.2 | 126,735 | 30 | 219,115 | 80 | 32,852 | 2.5 | 126,745 | 35 | 1.11 (0.79-1.66) | 1.11 (0.79-1.58) | 1.23 (0.87-1.76) | 1.19 (0.84-1.69) | 1.19 (0.74-1.93) |
| 127-182 | 148,040 | 40 | 20,484 | 2.0 | 121,960 | 35 | 206,665 | 55 | 27,796 | 2.0 | 121,740 | 25 | 1.03 (0.66-1.56) | 1.03 (0.69-1.55) | 1.11 (0.74-1.68) | 1.09 (0.72-1.64) | 0.76 (0.45-1.28) |
| **1-182** | **157,065** | **130** | **73,980** | **1.8** | **127,345** | **85** | **220,330** | **195** | **102,761** | **1.9** | **127,345** | **85** | **1.08 (0.86-1.36)** | **1.08 (0.86-1.35)** | **1.19 (0.95-1.49)** | **1.15 (0.92-1.45)** | **0.99 (0.73-1.34)** |
| **Non-COVID-19 death** | | | | | | | | | | | | | | | | | |
| 1-14 | 157,065 | 55 | 6,017 | 8.8 | 127,345 | 40 | 220,330 | 80 | 8,440 | 9.5 | 127,345 | 40 | 1.08 (0.73-1.49) | 1.08 (0.76-1.52) | 1.04 (0.73-1.47) | 1.07 (0.75-1.51) | 1.05 (0.68-1.64) |
| 15-70 | 156,900 | 425 | 24,007 | 17.8 | 127,225 | 310 | 220,070 | 570 | 33,674 | 17.0 | 127,210 | 300 | 0.95 (0.84-1.09) | 0.95 (0.84-1.08) | 0.93 (0.82-1.05) | 0.95 (0.83-1.07) | 0.96 (0.82-1.13) |
| 71-126 | 156,190 | 490 | 23,473 | 20.8 | 126,735 | 365 | 219,115 | 710 | 32,852 | 21.6 | 126,745 | 380 | 1.03 (0.92-1.16) | 1.03 (0.92-1.16) | 1.01 (0.90-1.13) | 1.03 (0.92-1.16) | 1.05 (0.91-1.21) |
| 127-182 | 148,040 | 490 | 20,484 | 23.9 | 121,960 | 355 | 206,665 | 600 | 27,796 | 21.6 | 121,740 | 330 | 0.91 (0.80-1.02) | 0.91 (0.80-1.02) | 0.89 (0.79-1.01) | 0.91 (0.81-1.03) | 0.92 (0.80-1.07) |
| **1-182** | **157,065** | **1,460** | **73,980** | **19.7** | **127,345** | **1,070** | **220,330** | **1,960** | **102,761** | **19.1** | **127,345** | **1,050** | **0.97 (0.90-1.03)** | **0.97 (0.91-1.04)** | **0.95 (0.88-1.01)** | **0.97 (0.90-1.04)** | **0.98 (0.90-1.07)** |

Counts are rounded to the nearest 5, and non-zero rounded counts of ≤10 redacted. AZ, AZD1222 (AstraZeneca); BNT, BNT162b2 (Pfizer/BioNTech); p-y, person-year; [R], redacted due to low event counts in one or both vaccine groups.

**Table S11. Incidence rates and hazard ratios for kidney disease subgroups (three-dose cohort).**

| | BNT–BNT–BNT | | | | AZ–AZ–BNT | | | | | |
|---|---|---|---|---|---|---|---|---|---|---|
| **Subgroup** | **N** | **N events** | **Person-years** | **Rate (per 1,000 p-y)** | **N** | **N events** | **Person-years** | **Rate (per 1,000 p-y)** | **Incidence rate ratio (95% CI)** | **Fully adjusted hazard ratio (95% CI)** |
| **Positive SARS-CoV-2 test** | | | | | | | | | | |
| **All** | **157,065** | **11,860** | **71,290** | **166.4** | **220,330** | **15,460** | **98,986** | **156.2** | **0.94 (0.92-0.96)** | **0.96 (0.93-0.98)** |
| CKD3 | 145,815 | 10,530 | 66,721 | 157.8 | 204,500 | 13,660 | 92,643 | 147.5 | 0.93 (0.91-0.96) | 0.95 (0.93-0.98) |
| CKD4–5 | 6,885 | 560 | 3,037 | 184.4 | 9,605 | 725 | 4,200 | 172.6 | 0.94 (0.84-1.05) | 0.96 (0.86-1.07) |
| RRT (transplant) | 2,885 | 480 | 965 | 497.6 | 4,445 | 715 | 1,471 | 486.0 | 0.98 (0.87-1.10) | 0.99 (0.88-1.11) |
| RRT (dialysis) | 1,480 | 290 | 568 | 510.8 | 1,785 | 355 | 672 | 528.5 | 1.03 (0.88-1.21) | 1.05 (0.89-1.23) |
| RRT (combined) | 4,365 | 765 | 1,532 | 499.2 | 6,230 | 1,075 | 2,143 | 501.7 | 1.00 (0.92-1.10) | 1.03 (0.94-1.13) |
| **COVID-19-related hospitalisation** | | | | | | | | | | |
| **All** | **157,065** | **1,400** | **73,762** | **19.0** | **220,330** | **1,960** | **102,428** | **19.1** | **1.01 (0.94-1.08)** | **1.01 (0.95-1.09)** |
| CKD3 | 145,815 | 1,090 | 68,955 | 15.8 | 204,500 | 1,500 | 95,758 | 15.7 | 0.99 (0.92-1.07) | 1.00 (0.93-1.08) |
| CKD4–5 | 6,885 | 140 | 3,145 | 44.5 | 9,605 | 220 | 4,335 | 50.8 | 1.14 (0.92-1.42) | 1.08 (0.87-1.34) |
| RRT (transplant) | 2,885 | 105 | 1,045 | 100.5 | 4,445 | 160 | 1,596 | 100.3 | 1.00 (0.78-1.29) | 1.01 (0.79-1.3) |
| RRT (dialysis) | 1,480 | 70 | 617 | 113.4 | 1,785 | 80 | 740 | 108.1 | 0.95 (0.68-1.33) | 0.98 (0.7-1.36) |
| RRT (combined) | 4,365 | 175 | 1,662 | 105.3 | 6,230 | 245 | 2,335 | 104.9 | 1.00 (0.82-1.22) | 1.01 (0.82-1.23) |
| **COVID-19-related death** | | | | | | | | | | |
| **All** | **157,065** | **130** | **73,980** | **1.8** | **220,330** | **195** | **102,761** | **1.9** | **1.08 (0.86-1.36)** | **1.15 (0.92-1.45)** |
| CKD3 | 145,815 | 90 | 69,120 | 1.3 | 204,500 | 130 | 96,010 | 1.4 | 1.04 (0.79-1.38) | 1.15 (0.87-1.51) |
| CKD4–5 | 6,885 | 15 | 3,169 | 4.7 | 9,605 | 30 | 4,374 | 6.9 | 1.45 (0.76-2.90) | 1.56 (0.80-3.02) |
| RRT (transplant) | 2,885 | 20 | 1,063 | 18.8 | 4,445 | 30 | 1,624 | 18.5 | 0.98 (0.54-1.82) | 1.09 (0.61-1.95) |
| RRT (dialysis) | 1,480 | ≤10 | 628 | [R] | 1,785 | ≤10 | 754 | [R] | [R] | [R] |
| RRT (combined) | 4,365 | 25 | 1,691 | 14.8 | 6,230 | 35 | 2,378 | 14.7 | 1.00 (0.58-1.74) | 1.04 (0.63-1.72) |
| **Non-COVID-19 death** | | | | | | | | | | |
| **All** | **157,065** | **1,460** | **73,980** | **19.7** | **220,330** | **1,960** | **102,761** | **19.1** | **0.97 (0.90-1.03)** | **0.97 (0.90-1.04)** |
| CKD3 | 145,815 | 1,180 | 69,120 | 17.1 | 204,500 | 1,575 | 96,010 | 16.4 | 0.96 (0.89-1.04) | 0.95 (0.88-1.03) |
| CKD4–5 | 6,885 | 185 | 3,169 | 58.4 | 9,605 | 285 | 4,374 | 65.2 | 1.12 (0.92-1.35) | 1.11 (0.92-1.34) |
| RRT (transplant) | 2,885 | 25 | 1,063 | 23.5 | 4,445 | 35 | 1,624 | 21.6 | 0.92 (0.53-1.60) | 0.90 (0.53-1.54) |
| RRT (dialysis) | 1,480 | 70 | 628 | 111.4 | 1,785 | 70 | 754 | 92.8 | 0.83 (0.59-1.18) | 0.84 (0.59-1.19) |
| RRT (combined) | 4,365 | 90 | 1,691 | 53.2 | 6,230 | 100 | 2,378 | 42.1 | 0.79 (0.59-1.06) | 0.83 (0.62-1.11) |

Counts are rounded to the nearest 5, and non-zero rounded counts of ≤10 redacted. AZ, AZD1222 (AstraZeneca); BNT, BNT162b2 (Pfizer/BioNTech); CKD, chronic kidney disease; p-y, person-year; [R], redacted due to low event counts in one or both vaccine groups; RRT, renal replacement therapy.
